# Clinically meaningful combined improvements of sleep, physical activity, and nutrition (SPAN) in relation to major adverse cardiovascular events

**DOI:** 10.1101/2025.09.10.25335537

**Authors:** Nicholas A. Koemel, Raaj Kishore Biswas, Stephen J. Simpson, Leandro F. M. Rezende, Tian Wang, Adrian Bauman, David Raubenheimer, Margaret Allman-Farinelli, Peter A. Cistulli, Matthew N. Ahmadi, Emmanuel Stamatakis

## Abstract

**Background:** Sleep, physical activity, and nutrition (SPAN) are major modifiable risk factors for cardiovascular disease, yet the minimum and optimal combined improvements for prevention remain unknown. We examined the multi-behaviour associations of SPAN with risk of major adverse cardiovascular events (MACE) and its subtypes (myocardial infarction (MI), heart failure (HF), and stroke).

**Methods:** This prospective cohort analysis included 53,242 participants from the UK Biobank (median age: 63.0 years; 56.8% male) who wore activity trackers for 7 days and self-reported dietary data. Wearable-measured sleep (hours/day) and moderate to vigorous physical activity (MVPA; mins/day) were calculated using a machine learning-based algorithm. A 10-item diet quality score (DQS) assessed intake of vegetables, fruits, whole grains and refined grains, unprocessed and processed meats, fish, dairy, vegetable oils, and sugary beverages. Cox proportional hazards models were used to estimate hazard ratios (HR) for MACE risk across 27 joint tertile combinations of SPAN behaviours. We examined dose-response associations of SPAN with MACE using a composite score (from 0-100 points).

**Results:** Over the 8.0-year median follow-up time, 2,034 MACE events occurred, including 932 myocardial infarctions, 584 strokes, and 518 HF events. Compared to the combined SPAN referent group (lowest tertiles for all three), the optimal SPAN combination involving high sleep duration (8.0-9.4 hours/day), high MVPA (42-104 mins/day), and a DQS between 32.5 and 50.0 was associated with an HR of 0.43 (95%CI: 0.30, 0.62). Compared to the minimum SPAN score of 17.8, a median SPAN score of 52.8 was associated with a 41% lower risk of MACE (HR: 0.59; 0.49, 0.70). The median SPAN score corresponded to an HR of 0.53 (0.38, 0.75) for HF, 0.65 (0.50, 0.84) for MI, and 0.52 (0.38, 0.71) for stroke. A theoretical minimum combined improvement of an additional 11 min/day of sleep, 4.5 min/day MVPA, and 3 DQS (1/4 cup of vegetables per day) were associated with 10% lower MACE risk (HR: 0.90; 0.88, 0.94).

**Conclusions:** Modest theoretical improvements across SPAN behaviours were associated with clinically meaningful reductions in MACE and its subtypes. These findings support multi-behavioural CVD prevention trials testing the effectiveness of small improvements across multiple behaviours.

## INTRODUCTION

Cardiovascular disease (CVD) is the leading cause of death worldwide^1^, which contributes to an increasingly unsustainable burden on healthcare systems from major clinical events such as myocardial infarction, heart failure, and stroke^2^. Lifestyle behaviours, including sleep, physical activity, and nutrition (SPAN), are key modifiable risk factors that physiologically underpin the development of cardiovascular disease^3^. Both under- and oversleeping have been linked to metabolic disruption, including worsened glucose homeostasis, elevated blood pressure, and upregulation of inflammation^4^. Physical inactivity is also directly related to CVD risk, impairing endothelial function, insulin sensitivity, and blood pressure regulation^5–7^. Poor diet quality and excess caloric intake have also contributed to CVD risk^8^, in particular, contributing to the rising obesity epidemic and worsened cardiometabolic health which directly increases the risk of cardiovascular complications^9,10^.

Traditionally, these behaviours have been explored as individual fields or disciplinary siloes. However, these behaviours are uniquely interdependent and have bidirectional impacts on one another. For example, poor sleep can disrupt the normal neurotransmission of appetite hormones, influencing food selection and driving an increase in total energy intake^4^. Physical activity has also been bidirectionally associated with sleep, where physical activity may improve sleep quality, while poor sleep may limit functional capacity due to fatigue^11,12^. Diet quality is also interlinked, demonstrating a relationship with sleep onset and wake time as well as energy and regulation for normal physical activity. Despite several studies examining distinct pairs of these behaviours^13,14^, few studies have explored all three behaviours in combination and the clinically relevant doses needed for CVD prevention. A recent study exploring the combination of SPAN^15^ revealed unique synergies between the behaviours, suggesting that a minimum dose of an additional 15 minutes of sleep, 1.5 minutes of moderate to vigorous physical activity, and ½ serving of vegetables per day was associated with a clinically meaningful 10% lower risk of all-cause mortality.

Previous intensive lifestyle interventions^16–18^ and population-level studies^19–21^ have largely focused on achieving broad, self-reported lifestyle targets, such as completing 150-300 minutes per week MVPA, meeting general dietary quality goals, or substantial caloric restriction. These one-size-fits-all approaches may discourage some individuals and hinder the initiation and sustainability of behaviour change^22^. Integrating simultaneous change across all three SPAN behaviours also has significant public health advantages as it reduces the required changes needed in any one behaviour to achieve meaningful clinical improvements^15^, likely resulting in more achievable lifestyle recommendations. Emerging clinical practice and research^23–25^ is shifting toward high-resolution personalised combined lifestyle approaches, yet limited evidence exists of clinically meaningful lifestyle targets for the general population.

This study aims to address this gap by exploring the association between combined SPAN and major adverse cardiovascular events (MACE), and to identify the minimum and incremental doses necessary to achieve a clinically meaningful improvement in MACE risk.

## METHODS

### Study Population

We used data from the UK Biobank accelerometry sub-study^26^, part of a wider cohort study of 502,629 adults aged 40-69 who were recruited from 2006 to 2010. All participants completed informed consent, and ethical approval was obtained by the UK National Health Service (NHS) and National Research Ethics Service for the UK (No. 11/NW/0382). Additional details of the study sample and study design are provided in **Supplementary Methods 1**.

### Outcome Ascertainment

Due to the nature of rolling updates for data linkage, participants were followed through 30 November 2022, where the date and cause of death were identified using the data linkage program with the National Health Service (NHS), Digital of England and Wales and the NHS Central Register and National Records of Scotland. Inpatient hospitalisation data were provided by either the Hospital Episode Statistics for England, the Patient Episode Database for Wales, or the Scottish Morbidity Record for Scotland. Instances of MACE were defined as CVD death or incidence of ST-elevated or non-ST elevated myocardial infarction (International Classification of Diseases Version 10: I21, I23, I24, I25, I26, I30, I31, I33, I34, I35, I38, I42, I45, I46, I48), stroke (I60, I61, I63, I64, I67) and heart failure (I11, I13, I50, I51).^27–31^

### Assessment of SPAN behaviours

Sleep and physical activity information was derived from wrist accelerometry data. All accelerometers were calibrated and initialised to 100 Hz and participants were instructed to wear the device on their dominant wrist for 7 consecutive days^26^. Sleep was defined as the average daily duration of sleep (hours/day), calculated using a validated algorithm based on relative changes in wrist tilt angle between successive 5-second windows^32,33^. We used time in moderate to vigorous physical activity (MVPA) as the main physical activity indicator in these analyses, estimated using a validated two-stage machine learning schema^27,29,34–37^ that categorises activities into light, moderate, or vigorous physical activity in 10-second epochs^27,29,34–37^. Additional details regarding the development, validation and performance of this schema are provided in Supplementary **Methods 2**.

From the recorded FFQ dietary information, we calculated a previously established diet quality score (DQS) which emphasises a higher intake of vegetables, fruits, fish, dairy, whole grains, and vegetable oils and a lower intake of refined grains, processed meats, unprocessed red meats, and sugar-sweetened beverages (**Supplemental Table 1**)^15,38^. This score rates the intake of each category on a scale from 0 (unhealthiest) to 10 (healthiest) for a total of 100 points, where higher values equate to a higher diet quality.

### Analysis of combined sleep, physical activity and nutrition

As per our previous work^15,39^, we first created 27 mutually exclusive groups of combined SPAN exposure (i.e., low, medium, high). The specific ranges for each exposure included sleep duration as 4.9–7.2 h/day (low), 7.2–8.0 h/day (medium), and 8.0–9.4 h/day (high); MVPA measurements as 5–23 min/day (low), 23–43 min/day (medium), and 43–103 min/day (high); and diet quality using the DQS as 32.5–50.0 (low), 50.0–57.5 (medium), and 57.5–72.5 (high). We used Cox proportional hazards models to assess the joint associations with MACE risk^27,30^. To account for competing risks from non-cardiovascular events, we employed a Fine-Gray sub-distribution hazards model^27,29,30,40^. We visualised the multivariable-adjusted associations of SPAN exposures with MACE by plotting each of the 27 categories as a forest plot with the lowest tertile of each exposure used as the reference point (**Figure 1**). Models were adjusted for age, sex, ethnicity, smoking, education, Townsend deprivation index, alcohol, discretionary screen time (time spent watching TV or using the computer outside of work), light intensity physical activity, medication (blood pressure, insulin, and cholesterol), previous diagnosis of major CVD (defined as disease of the circulatory system, arteries, and lymph, excluding hypertension), previous diagnosis of cancer, and familial history of CVD and cancer^15^. Further details on the covariates are provided in **Supplemental Table 2**. All models satisfied proportional hazards assumptions using Schoenfeld residuals. We examined the potential synergistic effects of SPAN behaviours using the relative excess risk due to interaction (RERI), attributable proportion due to interaction (AP), and the synergistic effects index (S)^28,41^ for MACE and its corresponding subtypes. These three indices estimate the contribution of synergistic interactions between exposures where an RERI or AP of 0 and an S value of 1 denote no interaction effect.

**Figure 1:**
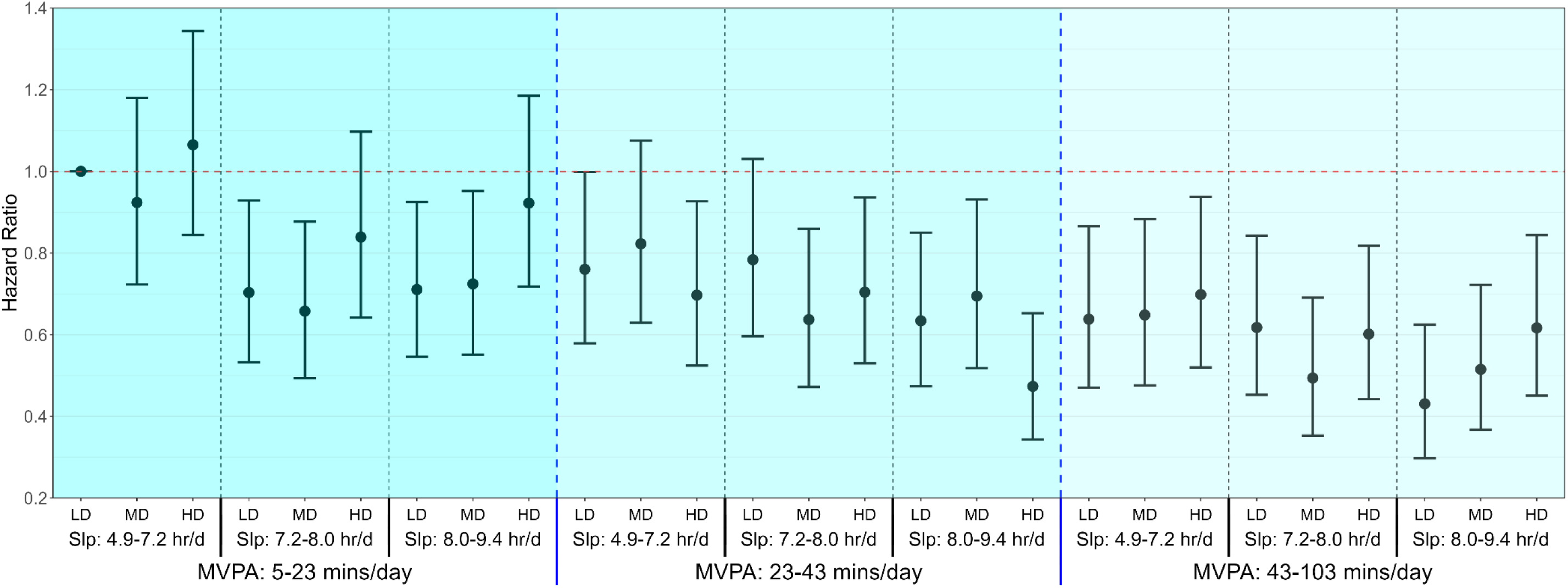
Multivariable-adjusted associations of combined sleep, physical activity, and nutrition with MACE risk (n = 53,242; events = 2,034) Model is adjusted for age, sex, ethnicity, smoking, education, Townsend deprivation index, alcohol, discretionary screen time (time spent watching TV or using the computer outside of work), light intensity physical activity, medication (blood pressure, insulin, and cholesterol), previous diagnosis of cancer, and familial history of CVD and cancer. Sleep (hours/day), physical activity (moderate to vigorous intensity (MVPA) minutes/day), and nutrition (Dietary Quality Score (DQS)) were included in the model as a joint term. Participants with a previous diagnosis of major CVD (defined as disease of the circulatory system, arteries, and lymph, excluding hypertension) were excluded from the analysis. The specific ranges for each exposure included sleep duration as 4.8-7.2 hours/day (low), 7·2-8.0 hours/day (medium), and 8.0-9.4 hours/day (high); MVPA measurements as 5-23 minutes/day (low), 23-42 minutes/day (medium), and 42-103 minutes/day (high); and diet quality using the DQS as 32.5-50.0 (low), 50.0-57.5 (medium), and 57.5-72.5 (high). The lowest tertiles for all three exposures (sleep, MVPA and DQS) was considered the reference group. Dashed blue lines separate tertiles MVPA and dashed black lines separate tertiles of sleep. Sleep (Slp); Low Diet Quality (LD); Medium Diet Quality (MD); High Diet Quality (HD).

### Continuous Sleep, Physical Activity, and Nutrition Score

We further examined the dose-response relationship between the combined and individual SPAN components using a continuous composite SPAN score (range: 0 to 100), with higher scores indicating healthier collective SPAN behaviours and each behaviour weighted equally. Higher scores reflect closer alignment to the theoretically optimal dose identified from the dose-response relationship with MACE^42,43^. For example, a U-shaped relationship was observed for sleep, whereby the nadir of the curve (i.e., theoretically most optimal amount of sleep) received the highest number of points, while under- or over-sleeping contributed a lower number of points to the SPAN score. We defined clinically meaningful as increments of 10% lower risk for MACE or its corresponding subtypes^15,44^. To further evaluate the clinically meaningful doses of SPAN with MACE risk, we created a heatmap correlogram that uses the 5^th^ percentile of all three exposures as a reference^15^, while providing the corresponding MACE risk associated with combined SPAN increments.

### Sensitivity Analyses

We completed a series of sensitivity analyses to examine how results vary by different analysis assumptions:

- To account for alternative sleep characteristics, we created a model that adjusted for self-reported insomnia, snoring, chronotype (morning/evening person), and daytime sleepiness^14^.
- To ensure findings were robust to the choice of dietary indicator, we repeated the primary analyses using the proportion of dietary ultra-processed food as a marker of diet quality (**Supplemental Table 3**)^45,46^.
- We also completed an analysis adjusting the model for total energy intake and excluded participants with sex-specific implausible ranges^47,48^.
- To minimise the influence of reverse causation we repeated the core analyses after excluding participants who self-reported poor health, current smokers, those in the top 20th percentile of the frailty index, and those with an underweight BMI (<18.5).
- We also repeated the main analysis excluding those with an event in the first two years of follow-up to assess potential reverse of causation^49^.
- To account for the potential mediating role of BMI^50–52^ in the associations between SPAN exposures and MACE, we repeated the main analyses adjusted for BMI.

We undertook all statistical analyses and visualisations using the survival, rms, ggplot2 packages of R (version 4.4.2). We followed the Strengthening the Reporting of Observational Studies in Epidemiology (STROBE) guidelines (**Supplemental Table 4**).

## RESULTS

### Sample

The final analytical sample in this study included 53,242 participants (median age [IQR]: 63 [56, 68] years; 56.8% male; **Supplementary Figure 1**). Over the 8.0-year follow-up period, 2034 MACE events occurred, comprising 932 myocardial infarctions, 584 strokes, and 518 heart failure events. We present baseline characteristics across tertiles of SPAN behaviours in **Table 1**.

**Table 1:** Distribution of SPAN exposures across participant characteristics.

|  | Overall | Sleep |  |  | Physical activity |  |  | Nutrition |  |  |
| --- | --- | --- | --- | --- | --- | --- | --- | --- | --- | --- |
|  |  | Low | Medium | High | Low | Moderate | High | Low | Moderate | High |
| **Sample** | 53,242 | 17,749 | 17,746 | 17,747 | 17,748 | 17,747 | 17,747 | 18,889 | 17,617 | 16,736 |
| **Events, n** | 2,034 (3.8%) | 829 (4.7%) | 608 (3.4%) | 597 (3.4%) | 914 (5.1%) | 639 (3.6%) | 481 (2.7%) | 692 (3.7%) | 630 (3.6%) | 712 (4.3%) |
| **Follow up, years (median [IQR])** | 8.04 [7.47, 8.56] | 8.04 [7.45, 8.55] | 8.06 [7.49, 8.58] | 8.04 [7.49, 8.58] | 8.04 [7.43, 8.59] | 8.04 [7.49, 8.57] | 8.06 [7.49, 8.55] | 8.05 [7.47, 8.57] | 8.05 [7.47, 8.59] | 8.03 [7.46, 8.55] |
| **Age, years (median [IQR])** | 63.00 [56.00, 68.00] | 64.00 [56.00, 69.00] | 63.00 [56.00, 68.00] | 64.00 [57.00, 68.00] | 65.00 [59.00, 70.00] | 63.00 [56.00, 68.00] | 61.00 [54.00, 67.00] | 62.00 [54.00, 67.00] | 64.00 [57.00, 68.00] | 65.00 [58.00, 69.00] |
| **Male, n (%)** | 30243 (56.8%) | 8283 (46.7%) | 7659 (43.2%) | 7057 (39.8%) | 7072 (39.8%) | 7762 (43.7%) | 8165 (46.0%) | 9172 (48.6%) | 6982 (39.6%) | 6845 (40.9%) |
| **Sleep, hours/day (median [IQR])** | 7.63 [6.90, 8.26] | 6.53 [5.95, 6.90] | 7.63 [7.42, 7.83] | 8.52 [8.26, 8.89] | 7.60 [6.80, 8.30] | 7.66 [6.94, 8.28] | 7.62 [6.96, 8.22] | 7.61 [6.88, 8.25] | 7.63 [6.92, 8.26] | 7.65 [6.91, 8.28] |
| **Moderate to vigorous physical activity, mins/day (median [IQR])** | 32.04 [19.14, 49.93] | 31.07 [18.26, 49.05] | 33.81 [20.38, 52.31] | 31.29 [18.86, 48.43] | 14.93 [10.19, 19.14] | 32.05 [27.53, 37.12] | 59.67 [49.93, 75.86] | 31.41 [18.95, 49.05] | 32.31 [19.26, 50.17] | 32.57 [19.24, 50.69] |
| **Diet quality score (median [IQR])** | 54.01 [47.50, 60.00] | 53.93 [47.50, 60.00] | 53.93 [47.50, 60.00] | 54.64 [47.50, 60.00] | 53.93 [47.50, 60.00] | 53.93 [47.50, 60.00] | 54.37 [47.50, 60.00] | 45.00 [40.00, 47.50] | 55.00 [52.50, 56.79] | 62.50 [60.00, 67.50] |
| **Light physical activity, mins/day (median [IQR])** | 103.54 [70.67, 158.91] | 107.48 [72.88, 163.39] | 105.91 [72.31, 162.76] | 97.29 [67.24, 150.34] | 84.86 [60.69, 119.84] | 108.15 [73.55, 158.40] | 131.39 [81.79, 196.71] | 101.57 [69.55, 156.31] | 104.43 [71.69, 159.39] | 104.67 [70.95, 160.73] |
| **Discretionary screen time, hours/day (median [IQR])** | 3.50 [2.50, 5.00] | 3.50 [2.50, 5.00] | 3.50 [2.50, 5.00] | 3.50 [2.50, 5.00] | 4.00 [2.50, 5.00] | 3.50 [2.50, 5.00] | 3.00 [2.00, 4.50] | 3.50 [2.50, 5.00] | 3.50 [2.50, 5.00] | 3.50 [2.50, 5.00] |
| **Smoking history, n (%)** |  | — | — | — | — | — | — | — | — | — |
| **Never** | 31,199 (58.6%) | 10,044 (56.6%) | 10,502 (59.2%) | 10,653 (60.0%) | 9,970 (56.2%) | 10,515 (59.2%) | 10,714 (60.4%) | 10,972 (58.1%) | 10,444 (59.3%) | 9,783 (58.5%) |
| **Former** | 18,747 (35.2%) | 6,394 (36.0%) | 6,199 (34.9%) | 6,154 (34.7%) | 6,428 (36.2%) | 6,211 (35.0%) | 6,108 (34.4%) | 6,358 (33.7%) | 6,239 (35.4%) | 6,150 (36.7%) |
| **Current** | 3,296 (6.2%) | 1,311 (7.4%) | 1,045 (5.9%) | 940 (5.3%) | 1,350 (7.6%) | 1,021 (5.8%) | 925 (5.2%) | 1,559 (8.3%) | 934 (5.3%) | 803 (4.8%) |
| **Alcohol consumption, units/week<sup>b</sup> (median [IQR])** | 9.75 [1.81, 19.50] | 9.50 [1.14, 19.50] | 9.75 [2.50, 19.50] | 9.75 [2.27, 19.50] | 8.77 [0.81, 18.54] | 9.75 [2.31, 19.50] | 10.42 [3.17, 20.42] | 9.75 [1.31, 19.80] | 9.75 [2.50, 19.50] | 9.75 [1.95, 19.50] |
| **Total energy intake, kcal/day (median [IQR])<sup>a</sup>** | 2,018 [1,651, 2,443] | 2,056 [1,676, 2,500] | 2,026 [1,662, 2,441] | 1,975 [1,617, 2,392] | 1,965 [1,607, 2,382] | 2,011 [1,649, 2,435] | 2,075 [1,699, 2,511] | 2,045 [1,677, 2,490] | 2,017 [1,652, 2,433] | 1,989 [1,620, 2,413] |
| **Townsend Deprivation Index, (median [IQR])** | -2.50 [-3.84, -0.26] | -2.33 [-3.76, 0.13] | -2.50 [-3.84, -0.33] | -2.64 [-3.92, -0.60] | -2.41 [-3.78, -0.14] | -2.54 [-3.88, -0.32] | -2.52 [-3.85, -0.32] | -2.44 [-3.82, -0.10] | -2.53 [-3.86, -0.39] | -2.52 [-3.85, -0.33] |
| **Education, n (%)** | — | — | — | — | — | — | — | — | — | — |
| **College/University** | 23,536 (44.2%) | 7,953 (44.8%) | 8,144 (45.9%) | 7,439 (41.9%) | 7,512 (42.3%) | 8,015 (45.2%) | 8,009 (45.1%) | 7,933 (42.0%) | 8,062 (45.8%) | 7,541 (45.1%) |
| **A/AS** | 7,064 (13.3%) | 2,397 (13.5%) | 2,307 (13.0%) | 2,360 (13.3%) | 2,326 (13.1%) | 2,392 (13.5%) | 2,346 (13.2%) | 2,502 (13.2%) | 2,446 (13.9%) | 2,116 (12.6%) |
| **O levels** | 10,798 (20.3%) | 3,494 (19.7%) | 3,512 (19.8%) | 3,792 (21.4%) | 3,690 (20.8%) | 3,562 (20.1%) | 3,546 (20.0%) | 3,983 (21.1%) | 3,515 (20.0%) | 3,300 (19.7%) |
| **CSE** | 2,042 (3.8%)<br>2042 (3.8%) | 665 (3.7%) | 662 (3.7%) | 715 (4.0%) | 583 (3.3%) | 635 (3.6%) | 824 (4.6%) | 859 (4.5%) | 618 (3.5%) | 565 (3.4%) |
| **NVQ/HND/HNC** | 2,801 (5.3%) | 931 (5.2%) | 925 (5.2%) | 945 (5.3%) | 961 (5.4%) | 953 (5.4%) | 887 (5.0%) | 1,084 (5.7%) | 856 (4.9%) | 861 (5.1%) |
| **Other** | 7,001 (13.1%) | 2,309 (13.0%) | 2,196 (12.4%) | 2,496 (14.1%) | 2,676 (15.1%) | 2,190 (12.3%) | 2,135 (12.0%) | 2,528 (13.4%) | 2,120 (12.0%) | 2,353 (14.1%) |
| **Parental history of CVD, n (%)** | 29,287 (55.0%) | 9,767 (55.0%) | 9,707 (54.7%) | 9,813 (55.3%) | 10,098 (56.9%) | 9,759 (55.0%) | 9,430 (53.1%) | 9,860 (52.2%) | 9,815 (55.7%) | 9,612 (57.4%) |
| **Parental history of cancer, n (%)** | 16,797 (31.5%) | 5,608 (31.6%) | 5,560 (31.3%) | 5,629 (31.7%) | 5,624 (31.7%) | 5,651 (31.8%) | 5,522 (31.1%) | 5,902 (31.2%) | 5,618 (31.9%) | 5,277 (31.5%) |
| **Previous Cancer, n (%)** | 4,365 (8.2%) | 16,336 (92.0%) | 16,312 (91.9%) | 16,229 (91.4%) | 17,09 (9.6%) | 1,460 (8.2%) | 1,196 (6.7%) | 1,440 (7.6%) | 1,505 (8.5%) | 1,420 (8.5%) |
| **Ethnicity, n (%)** | — | — | — | — | — | — | — | — | — | — |
| **Other** | 3,290 (6.2%) | 1,433 (8.1%) | 1,026 (5.8%) | 831 (4.7%) | 1,061 (6.0%) | 1,083 (6.1%) | 1,146 (6.5%) | 1,166 (6.2%) | 1,085 (6.2%) | 1,039 (6.2%) |
| **White** | 49,952<br>(93.8%) | 16,316<br>(91.9%) | 16,720<br>(94.2%) | 16,916<br>(95.3%) | 16,687<br>(94.0%) | 16,664<br>(93.9%) | 16,601<br>(93.5%) | 17,723<br>(93.8%) | 16,532<br>(93.8%) | 15,697<br>(93.8%) |
| **Medication Use, n (%)** | — | — | — | — | — | — | — | — | — | — |
| **Cholesterol** | 6,442<br>(12.1%) | 2,410<br>(13.6%) | 1,969<br>(11.1%) | 2,063<br>(11.6%) | 2,884<br>(16.2%) | 2,056<br>(11.6%) | 1,502 (8.5%) | 1,963<br>(10.4%) | 2,044<br>(11.6%) | 2,435<br>(14.5%) |
| **Blood pressure** | 4,637 (8.7%) | 1,702 (9.6%) | 1,448 (8.2%) | 1,487 (8.4%) | 1,991<br>(11.2%) | 1,500 (8.5%) | 1,146 (6.5%) | 1,523 (8.1%) | 1,500 (8.5%) | 1,614 (9.6%) |
| **Insulin** | 64 (0.1%) | 17 (0.1%) | 27 (0.2%) | 20 (0.1%) | 22 (0.1%) | 25 (0.1%) | 17 (0.1%) | 20 (0.1%) | 16 (0.1%) | 28 (0.2%) |
| **Frailty Index >3, n (%)** | 148 (0.3%) | 68 (0.4%) | 39 (0.2%) | 41 (0.2%) | 105 (0.6%) | 20 (0.1%) | 23 (0.1%) | 69 (0.4%) | 43 (0.3%) | 36 (0.2%) |
| **Self-rated health, n (%)** | — | — | — | — | — | — | — | — | — | — |
| **Excellent** | 11,821<br>(23.3%) | 10,082<br>(59.6%) | 10,388<br>(61.3%) | 4,040<br>(23.9%) | 10,273<br>(60.7%) | 4,065<br>(24.0%) | 4,775<br>(28.2%) | 10,802<br>(60.1%) | 3,999<br>(23.8%) | 3,992<br>(25.0%) |
| **Good** | 30,893<br>(60.8%) | 2,769<br>(16.4%) | 2,146<br>(12.7%) | 10,423<br>(61.6%) | 3,138<br>(18.5%) | 10,457<br>(61.8%) | 10,163<br>(60.0%) | 2,871<br>(16.0%) | 10,373<br>(61.6%) | 9,718<br>(60.8%) |
| **Fair** | 7,080<br>(13.9%) | 401 (2.4%) | 250 (1.5%) | 2,165<br>(12.8%) | 508 (3.0%) | 2,136<br>(12.6%) | 1,806<br>(10.7%) | 430 (2.4%) | 2,186<br>(13.0%) | 2,023<br>(12.6%) |
| **Poor** | 927 (1.8%) | 26 (0.2%) | 20 (0.1%) | 276 (1.6%) | 29 (0.2%) | 252 (1.5%) | 167 (1.0%) | 26 (0.1%) | 248 (1.5%) | 249 (1.6%) |
| **Body mass index, kg/m<sup>2</sup>**<br>**(median [IQR])** | 25.90<br>[23.50,<br>28.80] | 26.50<br>[24.00,<br>29.60] | 25.70<br>[23.40,<br>28.60] | 25.50<br>[23.20,<br>28.30] | 27.00<br>[24.40,<br>30.30] | 25.80<br>[23.50,<br>28.60] | 25.10<br>[22.90,<br>27.50] | 26.20<br>[23.80,<br>29.30] | 25.80<br>[23.50,<br>28.70] | 25.70<br>[23.30,<br>28.50] |

### Combined SPAN associations with MACE risk

In the overall gradient of behaviours, MVPA contributed the most to the association with MACE, followed by sleep and diet (**Figure 1**). Compared to the referent category (theoretically least healthy of all three behaviours), low MVPA, moderate sleep and moderate diet were associated with a 34% lower MACE risk (HR: 0.66; 95% CI: 0.49, 0.88). Moderate MVPA, moderate sleep, and moderate DQS were associated with 36% lower risk of MACE (HR: 0.64; 95% CI: 0.47, 0.86). The lowest risk group associated with MACE was the high MVPA, high sleep, and low DQS (HR: 0.43; 95% CI: 0.30, 0.62), although a similar level of risk was observed in the moderate MVPA, high sleep, and high DQS category (HR: 0.47; 95% CI: 0.34, 0.62).

### Synergistic relationship of SPAN behaviours with MACE

We observed a non-statistically significant positive synergistic interaction between SPAN behaviours for overall MACE (RERI = 0.003; 95% CI: −0.03, 0.04; AP = 0.4%; 95% CI: − 6-7%; S = 1.03; 95% CI: −3.32, 5.29; **Supplementary Figure 2**). Similar non-significant results were found for HF (RERI = −0.02; 95% CI: −0.11, 0.07; AP = −2.6%; 95% CI: −24-18%; S = 1.07; 95% CI: −4.46, 6.59) and MI (RERI = −0.003; 95% CI: −0.05, 0.04; AP = −0.3%; 95% CI: −8-8%; S = 1.03; 95% CI: 0.18, 1.89). We observed a weaker, non-statistically significant positive synergistic interaction for stroke (RERI = 0.038; 95% CI: - 0.07, 0.15; AP = 6%; 95% CI: −40-51%; S = 0.90; 95% CI: 0.57, 1.22).

### Dose-response relationship of SPAN behaviours with MACE risk

The dose-response relationship for the individual SPAN behaviours with MACE and the corresponding subtypes is shown in **Supplementary Figure 3**. Sleep displayed an L-shaped relationship for MACE, MI, and stroke, while a linear dose-response relationship was observed for HF. We observed an L-shaped relationship with physical activity for all outcomes, while nutrition displayed a subtle, non-statistically significant association.

We present the dose-response associations of the combined continuous composite SPAN score with MACE, MI, HF, and stroke risk in **Figure 2**. There was a near-linear inverse relationship between SPAN score and MACE risk, wherein compared to the 5^th^ percentile of the SPAN score, the median (52.5) and maximum SPAN score (85.2), were associated with a 41% (HR: 0.59; 95% CI: 0.49, 0.70) and 50% lower risk (HR: 0.50; 95% CI: 0.40, 0.62) of MACE, respectively. We found a linear relationship of the SPAN score with MI and HF, with median values corresponding to an HR of 0.54 (95% CI: 0.38, 0.75) and 0.65 (95% CI: 0.50, 0.84), respectively. There was an L-shaped relationship between SPAN score and stroke risk, with a median SPAN score associated with an HR of 0.52 (95% CI: 0.38, 0.71).

**Figure 2:**
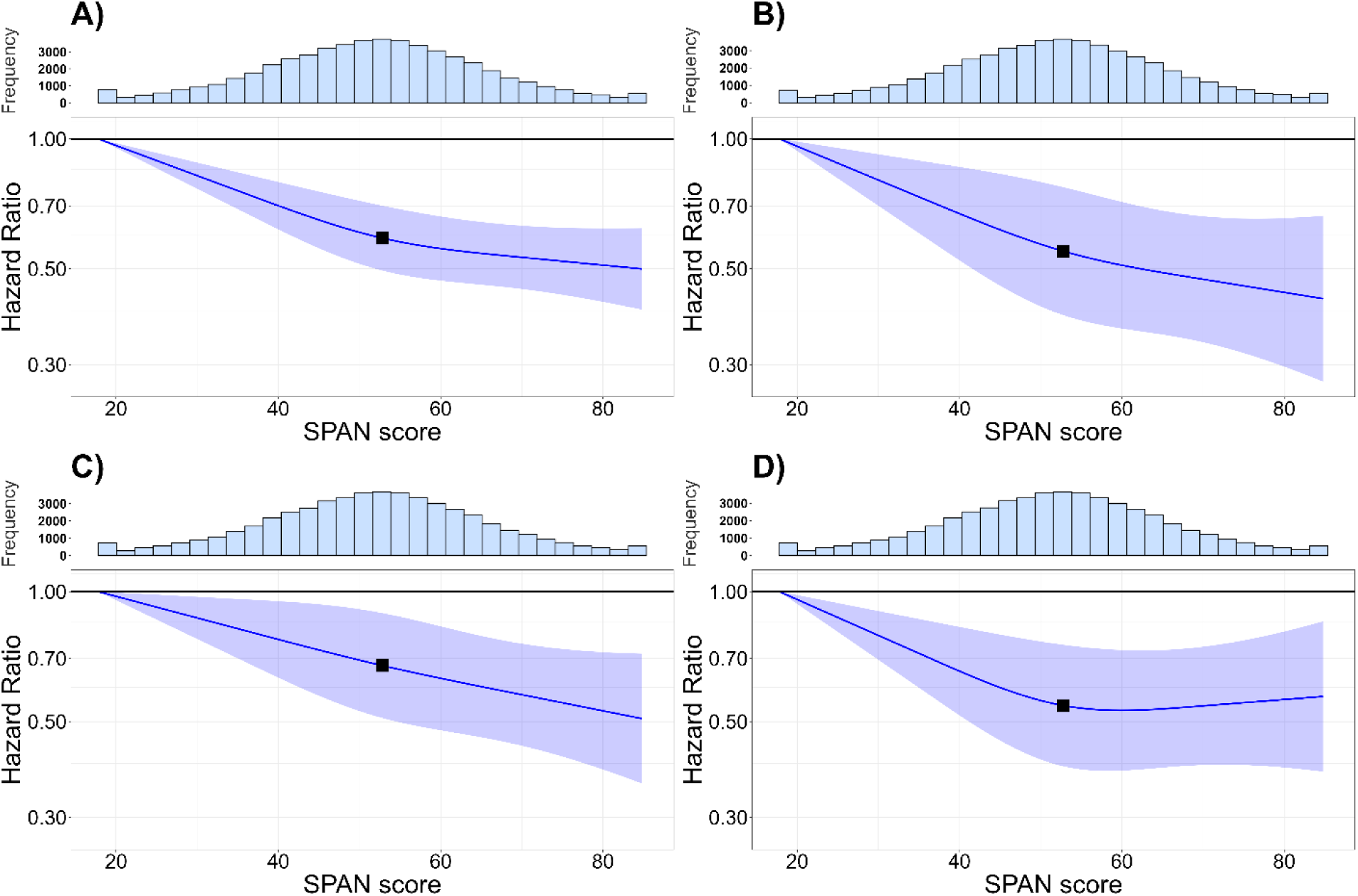
Multivariable-adjusted associations of combined sleep, physical activity, and nutrition with risk of A) MACE, B) heart failure, C) myocardial infarction, and D) stroke (n = 53,242; events = 2,034) Model is adjusted for age, sex, ethnicity, smoking, education, Townsend deprivation index, alcohol, discretionary screen time (time spent watching TV or using the computer outside of work), light intensity physical activity, medication (blood pressure, insulin, and cholesterol), previous diagnosis of cancer, and familial history of CVD and cancer. Participants with a previous diagnosis of major CVD (defined as disease of the circulatory system, arteries, and lymph, excluding hypertension) were excluded from the analysis. A combined SPAN score ranging from 0 to 100 was developed by assigning equal weights (33.33 points) to three lifestyle components: Sleep (hours/day), physical activity (moderate to vigorous physical activity (MVPA) minutes/day), and diet quality (Dietary Quality Score, DQS). Scoring for each component was based on the observed sample range. For physical activity, participants with the lowest MVPA received a score of 0, while those with the highest MVPA scored 33.33, with intermediate values scaled proportionally. Diet quality was scored similarly, from 0 for the lowest DQS to 33.33 for the highest. However, because sleep duration had a U-shaped association with MACE, a score of 33.33 was assigned to the optimal sleep duration of 7.9 hours/day. Sleep durations above and below 7.9 hours were scored proportionally lower, tapering symmetrically on either side of this nadir.

### Clinically meaningful combined thresholds of SPAN behaviours associated with MACE risk

We present the minimum and incremental clinically meaningful doses (10-60%) of combined SPAN behaviours for MACE, HF, MI, and stroke risk in **Table 2**. The rows indicate doses of both combined and individual SPAN behaviours associated with increments of 10% lower risk for each outcome. Compared to the 5^th^ percentile of sleep (5.5 hours per day), physical activity (7.3 minutes per day of MVPA), and diet (36.9 DQS points), a combined improvement of 11 minutes/day of sleep, 4.5 minutes of MVPA, and 3 DQS points was associated with a 10% lower risk of MACE. A similar minimum dose of combined SPAN behaviours was observed for HF (additional 9 min/day sleep, 3.7 min/day MVPA, 2 DQS; HR: 0.91; 95%CI: 0.86, 0.96), MI (additional 15 min/day sleep, 6 min/day MVPA, and 4 DQS; HR: 0.90; 95%CI: 0.84, 0.97), and stroke (additional 9 min/day sleep, 4.9 min/day MVPA, and 2 DQS; HR: 0.90; 95%CI: 0.86, 0.95). Higher doses, such as 62 minutes of additional sleep per day, 24.6 minutes per day of MVPA, and 16 additional DQS points, were associated with a 40% lower risk of MACE (HR: 0.60; 95% CI: 0.50, 0.71). We provide an additional visualisation of combined SPAN behaviour doses and corresponding MACE risk as a heatmap in **Figure 3** (MACE subtypes are presented in **Supplementary Figures 4-6**).

**Figure 3:**
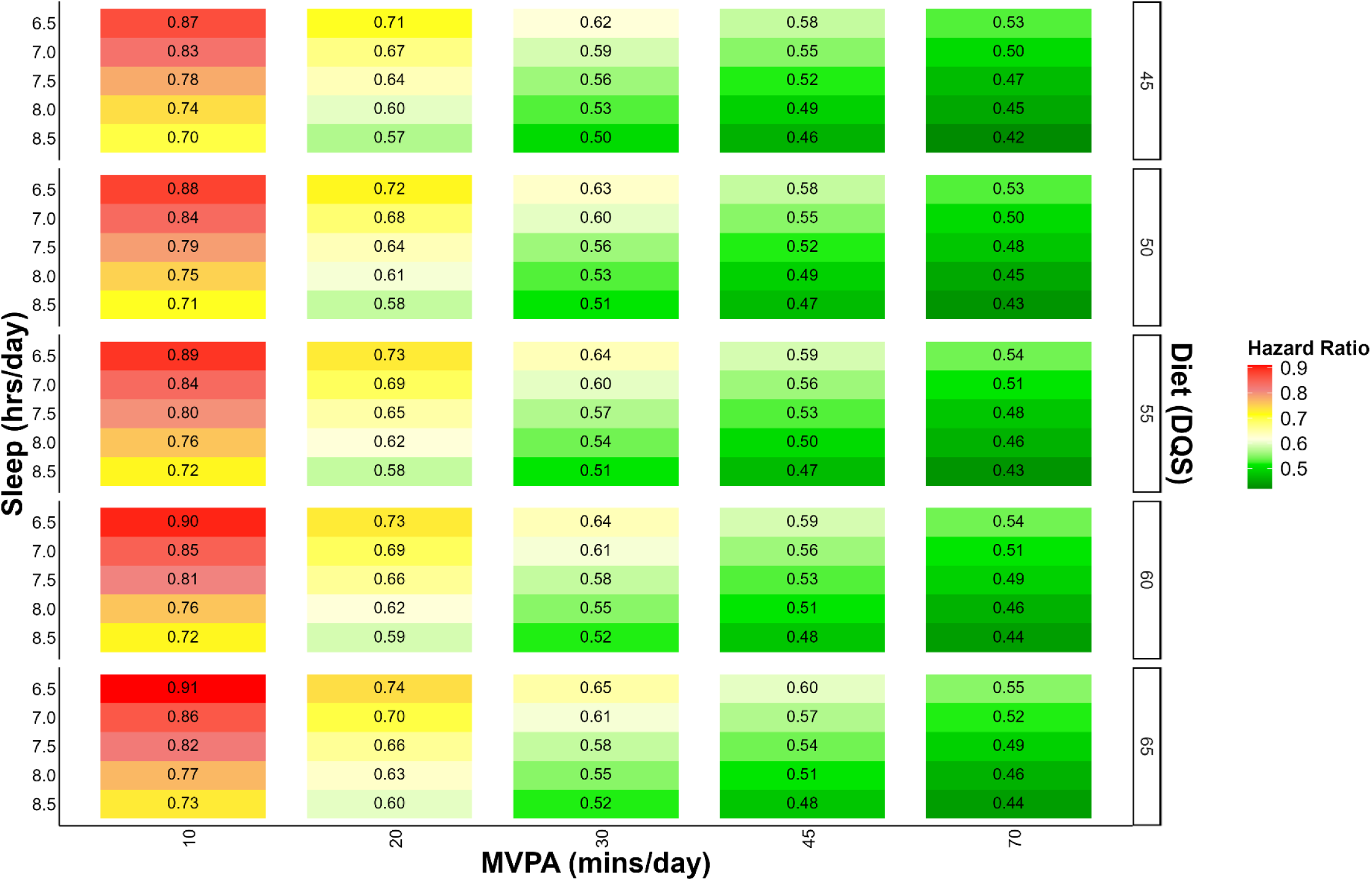
Multivariable adjusted MACE risk associated with concurrent variations in sleep, physical activity, and nutrition (n = 53,242; events = 2,034) The correlogram displays changes in sleep (hours/day), physical activity (moderate to vigorous intensity (MVPA) minutes/day), and nutrition (Dietary Quality Score (DQS)) and corresponding MACE risk with the reference being the 5^th^ percentile of sleep (5.5 hours/day), physical activity (7.9 minutes/day), and nutrition (37.2 DQS). Sleep, physical activity, and nutrition are included as independent terms in the model to allow for more granular predictions. Each square on the grid represents the hazard ratio for MACE associated with a combination of behaviours, as defined by the x-axis (physical activity), y-axis (sleep), and z-axis (nutrition). The colour corresponds to the hazard ratio where red indicates a higher risk of MACE and green indicates a lower risk of MACE. Model is adjusted for age, sex, ethnicity, smoking, education, Townsend deprivation index, alcohol, discretionary screen time (time spent watching TV or using the computer outside of work), light intensity physical activity, medication (blood pressure, insulin, and cholesterol), previous diagnosis of cancer, and familial history of CVD and cancer. Participants with a previous diagnosis of major CVD (defined as disease of the circulatory system, arteries, and lymph, excluding hypertension) were excluded from the analysis.

**Table 2:** Clinically meaningful combined improvements of sleep, physical activity, and nutrition associated with MACE, heart failure, myocardial infarction, and stroke risk. **Table 2** displays the clinically meaningful concurrent doses of sleep, physical activity and nutrition associated with MACE risk compared to individual SPAN exposures. The combined columns show the SPAN combinations and corresponding MACE risk compared to the 5th percentile of sleep (5.5 hours/day), moderate to vigorous physical activity (7.3 minutes/day), nutrition (36.9 DQS), and composite SPAN score (17.8) in increments of 10%. For comparison, the dose needed for individual SPAN exposures is shown on the right (greyed cells). Empty cells denote that the individual SPAN exposure could not achieve that level of risk reduction in isolation. All models were adjusted for age, sex, ethnicity, smoking, education, Townsend deprivation index, alcohol, discretionary screen time (time spent watching TV or using the computer outside of work), light intensity physical activity, medication (blood pressure, insulin, and cholesterol), previous diagnosis of cancer, and familial history of CVD and cancer. Participants with a previous diagnosis of major CVD (defined as disease of the circulatory system, arteries, and lymph, excluding hypertension) were excluded from the analysis. Moderate to vigorous physical activity (MVPA); Diet quality score (DQS); Hazard ratio (HR). Each exposure was adjusted by the median value of the other two SPAN exposures. *Values were non-significant. ^†^95% CI are based on for combined SPAN model.

| Clinically meaningful increments of risk<sup>†</sup> | Additional SPAN Score | Additional Sleep (min/day) | Additional Moderate to Vigorous Physical Activity (min/day) | Additional Nutrition (DQS points) | Additional Sleep* (min/day) | Additional Moderate to Vigorous Physical Activity (min/day) | Additional Nutrition (DQS points) |
| --- | --- | --- | --- | --- | --- | --- | --- |
| **MACE** |  | **Combined SPAN** |  |  | **Individual SPAN** |  |  |
| 10% lower risk<br>(HR: 0.91; 95%CI: 0.88, 0.94) | 6 | 11 | 4.5 | 3 | 30 | 6.6 | 15* |
| 20% lower risk<br>(HR: 0.80; 95%CI: 0.74, 0.86) | 14 | 26 | 10.5 | 7 | 66 | 14 | - |
| 30% lower risk<br>(HR: 0.70; 95%CI: 0.58, 0.84) | 22 | 41 | 16.4 | 11 | 174 | 24.6 | - |
| 40% lower risk<br>(HR: 0.60; 95%CI: 0.50, 0.71) | 33 | 62 | 24.6 | 16 | - | 50.6 | - |
| 50% lower risk<br>(HR: 0.50; 95%CI: 0.41, 0.62) | 63 | 119 | 47.8 | 30 | - | - | - |
| **Heart Failure** |  | **Combined SPAN** |  |  | **Individual SPAN** |  |  |
| 10% lower risk<br>(HR: 0.91; 95%CI: 0.86, 0.96) | 5 | 9 | 3.7 | 2 | 32 | 4.5 | 13* |
| 20% lower risk<br>(HR: 0.80; 95%CI: 0.70, 0.91) | 12 | 22 | 9.0 | 6 | 62 | 10.0 | - |
| 30% lower risk<br>(HR: 0.70; 95%CI: 0.57, 0.86) | 19 | 35 | 14.2 | 9 | 104 | 16.0 | - |
| 40% lower risk<br>(HR: 0.60; 95%CI: 0.45, 0.81) | 27 | 50 | 20.2 | 13 | 182 | 25.0 | - |
| 50% lower risk<br>(HR: 0.50; 95%CI: 0.36, 0.70) | 41 | 76 | 30.6 | 20 | - | 54.0 | - |
| 60% lower risk<br>(HR: 0.42; 95%CI: 0.28, 0.65) | 67 | 125 | 50.02 | 32 | - | - | - |

| Myocardial Infarction |  | Combined SPAN |  |  | Individual SPAN |  |  |
| --- | --- | --- | --- | --- | --- | --- | --- |
| 10% lower risk<br>(HR: 0.90; 95%CI: 0.84, 0.97) | 8 | 15 | 6.0 | 4 | 49 | 6.0 | 16* |
| 20% lower risk<br>(HR: 0.80; 95%CI: 0.68, 0.93) | 18 | 34 | 13.4 | 9 | 103 | 12.5 |  |
| 30% lower risk<br>(HR: 0.70; 95%CI: 0.56, 0.89) | 28 | 52 | 20.9 | 13 | - | 21.0 | - |
| 40% lower risk<br>(HR: 0.60; 95%CI: 0.47, 0.78) | 43 | 80 | 32.1 | 20 | - | 36.5 | - |
| 50% lower risk<br>(HR: 0.50; 95%CI: 0.37, 0.69) | 66 | 123 | 49.3 | 31 | - | - | - |
| Stroke |  | Combined SPAN |  |  | Individual SPAN |  |  |
| 10% lower risk<br>(HR: 0.90; 95%CI: 0.86, 0.95) | 5 | 9 | 4.9 | 2 | 26 | 13.0 | 7.0* |
| 20% lower risk<br>(HR: 0.80; 95%CI: 0.71, 0.89) | 11 | 21 | 10.8 | 5 | 50 | 30.5 | 16* |
| 30% lower risk<br>(HR: 0.70; 95%CI: 0.59, 0.84) | 17 | 32 | 16.7 | 8 | 80 | 71.5 | - |
| 40% lower risk<br>(HR: 0.60; 95%CI: 0.46, 0.78) | 25 | 46 | 24.5 | 11.9 | - | - | - |
| 50% lower risk<br>(HR: 0.50; 95%CI: 0.37, 0.71) | 38 | 70.8 | 37.2 | 18.1 | - | - | - |

### Sensitivity analyses

Excluding individuals who had poor health at baseline (n = 50,787; events = 1,907) or those with an event in the first two years of follow-up resulted in no major changes in the underlying findings of the study (**Supplementary Figure 7-8**). Additional adjustments for BMI (n = 52,744; events = 2,004) or additional sleep characteristics (n = 33,518; events = 1241) did not impact the results (**Supplementary Figure 9-10**). We also observed no materially different results when using the proportion of diet from ultra processed foods as an alternative dietary quality index (n = 39,533; events = 1,462) or adjusting for total energy intake (n = 38,921; events = 1,441; **Supplementary Figure 11-12**).

## DISCUSSION

### Main Findings

To date, this study presents the first investigation into clinically relevant combined doses of SPAN behaviours in relation to MACE risk. We apply a novel multi-behaviour framework^15^ and a large wearable sub-study from the UK Biobank to examine the association of combined behaviour variations with MACE and its subtypes. We show that when compared to the lowest tertile of all exposures, the optimal combinations of SPAN behaviours consisted of 8.0-9.4 hours per day of sleep, 42-104 minutes per day of MVPA, and a DQS score of 32.50-50.0, which was associated with a 57% lower risk of MACE 0.43 (95% CI: 0.30-0.62). We also identified clinically relevant improvements of the behaviours, which in combination yielded significantly lower MACE risk through relatively modest combined variations. For example, a minimum clinically relevant combined improvement of an additional 10 minutes per day of sleep, approximately 5 minutes per day of MVPA, and 3 DQS (equivalent to an extra 1/4 cup of vegetables), was associated with a 10% lower risk of MACE.

Previous studies^53–55^ involving lifestyle behaviours and cardiovascular disease risk have primarily used broad-level self-reported categories of behaviours, such as dichotomously categorising participants as healthy or unhealthy. Studies often provide general suggestions such that improving a combination of lifestyle behaviours, including alcohol, smoking, obesity, sleep, physical activity and diet, is associated with a 58-62% lower risk of cardiovascular disease^53–55^. However, this broad level of recommendation limits the ability to generate consumer-friendly, translatable public health messaging. For example, physical activity guidelines often recommend achieving 150 minutes per week of MVPA^56^, however, these recommendations lack the specificity needed to initiate and sustain behaviour change while taking into consideration other behaviours. Similar one-size-fits-all recommendations exist for sleep (e.g., optimal sleep of 7-8 hours per night for older adults)^57^ or the UK Eatwell Guide^58^, which recommends general serving food groups and portion goals. However, understanding the incremental variations needed for clinically meaningful improvements in cardiovascular risk is necessary for the implementation of more tailored and sustainable holistic lifestyle recommendations^24,59^.

Sleep, physical activity, and nutrition have been almost exclusively examined as individual behaviours or as joint pair-wise behaviours^13,14,60–63^. This narrow unidisciplinary approach fails to account for the many unique biological and behavioural interdependencies that exist between these behaviours^62,64–68^. For example, recent evidence^15^ suggests the unique collective synergies of these behaviours for the risk of all-cause mortality, whereby in combination, they offer clinically meaningful changes in risk with relatively modest combined variations. The present findings align with this previous work, suggesting that individual behaviours require substantially higher doses to achieve the same level of risk reduction. For example, we show that in isolation, sleep would require three times the amount (30 minutes) to achieve a 10% lower risk of MACE when compared to more modest amounts of sleep (≈10 minutes) in combination with physical activity (≈5 additional minutes MVPA per day) and nutrition (additional 1/4 cup of vegetables per day). These findings offer important insights into the feasibility of lifestyle changes, as making more substantial shifts in any one particular lifestyle behaviour may be less achievable and sustainable for some individuals.

Notably, we observed a unique heterogeneity for the SPAN relationship across MACE subtypes. The greatest overall benefit of SPAN was observed for HF with a linear dose-response relationship associated with 69% lower risk at the maximum observed SPAN score, compared with 49% for MI and 53% for stroke. Among the individual behaviours, sleep displayed the most pronounced differential relationship, whereby we observed a beneficial relationship with HF linearly compared to the L- and U-shaped relationship observed for MI and stroke, respectively. For all subtypes, increased risk from insufficient sleep is mechanistically supported by increased sympathetic nervous system activity, endothelial dysfunction, blood pressure, and inflammation^69^. Longer sleep periods may confer unique physiological benefits for HF through the restoration and maintenance of autonomic balance, the renin-angiotensin-aldosterone system, and blood pressure regulation^70^. In contrast, longer sleep for MI and stroke may be partially explained or exacerbate circadian dysregulation, which can increase pro-thrombotic triggering processes, including increased platelet aggregation, reduced fibrinolysis and abnormal blood pressure surges^71,72^. In line with these findings, there is a growing recognition of sleep as a relevant lifestyle characteristic, as evidenced by its recent addition to the American Heart Association’s Essential 8 behaviours for the prevention and management of cardiovascular disease^3^.

This study highlights the combined potential of SPAN behaviours in reducing MACE risk, with significant clinical and public health implications. For many individuals, meeting guideline-based targets, such as 150 minutes of MVPA per week, sufficient sleep, or recommended fruit and vegetable intake, can be challenging and is often met with feasibility concerns. Our results suggest that focusing on the combination of behaviours, rather than strict adherence to broad behavioural targets, may yield substantial health benefits while minimising the burden of change on any single behaviour. Moreover, by exploring the incremental improvements of SPAN needed for clinically meaningful shifts in MACE risk, these findings can help shape more holistic and behaviourally sustainable lifestyle recommendations with clear and translatable lifestyle targets, which is a fundamental tenet of sustainable behaviour change^24^.

### Strengths and limitations

A core strength of the study is a novel multi-behavioural analytical approach^15^ which enabled us to explore combined, clinically meaningful improvements of SPAN behaviours for MACE risk. Wearable-based metrics and machine learning approaches for these behaviours allowed us to explore incremental doses of these behaviours at a higher resolution than self-reported research, which typically characterises the behaviour in crude terms (i.e., >10-minute chunks) of physical activity^73^ and hourly increments of sleep^74^. For physical activity in particular, this resolution of wearables data offers a major advantage in this study, as the self-report surveys fail to capture the brief periods of movement that contribute to cardiovascular risk^27,29,73,75^.

However, a potential limitation of this study is the differing methods used to assess SPAN in this study. Sleep and physical activity were collected using wearable devices while nutrition was self-reported using a validated FFQ. This methodological discrepancy may partially explain the subtle, nonsignificant relationship between diet and MACE which differs from existing literature^8,76^. Dietary behaviours in the UK Biobank are also more significantly impacted by regression dilution bias^77^ which may have influenced its true relationship with MACE. In the UK Biobank^26,78^, there is also a median of 5.5 years lag between the data collection of self-reported dietary data and the wearable device measures of sleep and physical activity which could have impacted the underlying findings. Although we conducted a range of sensitivity analyses, including removing those of poor health and adjusting models for BMI, residual confounding cannot be ruled out. Further research is needed to explore the length of time under behaviour change needed to elicit health benefits and how sustainable these are across diverse populations. Lastly, although we explored the combined associations of sleep, physical activity, and nutrition, substance-use behaviours such as smoking and alcohol should be explored as a part of future studies as they also contribute to overall lifestyle patterns and cardiovascular risk.

## CONCLUSIONS

We show that even modest combined doses such as an additional 11 minutes per day of sleep, approximately 5 minutes per day of MVPA, and 1/4 cup of vegetables per day, were associated with a 10% lower risk of MACE. Additionally, we highlight that in combination, higher doses were able to achieve a significantly lower risk than the sum of the individual behaviours. This study supports that relatively modest theoretical improvements in SPAN behaviours are associated with clinically meaningful improvements in MACE risk. These findings highlight the need for integrated multi-behaviour lifestyle prevention trials to evaluate the effectiveness of small, achievable lifestyle changes for MACE prevention.

## Acknowledgements

This research has been conducted using the UK Biobank resource under application number 25813. The authors would like to thank all the participants and professionals contributing to the UK Biobank. All information and materials in the manuscript are original and have not been submitted for publication elsewhere.

## Data availability Statement

The data that support the findings of this study are available from the UK Biobank, but restrictions apply to the availability of these data, which were used under license for the current study, and so are not publicly available. Data are however available from the authors upon reasonable request and with the permission of the UK Biobank.

