## Supplementary material for "Clinically meaningful combined improvements of sleep, physical activity, and nutrition (SPAN) in relation to major adverse cardiovascular events": ONLINE SUPPLEMENTARY MATERIAL

| **Page** | **Item** |
| --- | --- |
| **3** | **Supplemental Figure 1:** Flow diagram of participants in the study |
| **4** | **Supplementary Figure 2:** Synergistic relationship between sleep, physical activity, and nutrition in relation to major adverse cardiovascular events |
| **5** | **Supplementary Figure 3:** Dose-response association of sleep, physical activity, and diet with MACE (A-C), heart failure (D-F), myocardial infarction (G-I), and stroke (J-L) |
| **7** | **Supplementary Figure 4:** Multivariable adjusted heart failure risk associated with concurrent variations in sleep, physical activity, and nutrition (n = 51,726; events = 518) |
| **8** | **Supplemental Figure 5:** Multivariable adjusted myocardial infarction risk associated with concurrent variations in sleep, physical activity, and nutrition (n = 52,140; events = 932) |
| **9** | **Supplementary Figure 6:** Multivariable adjusted stroke risk associated with concurrent variations in sleep, physical activity, and nutrition (n = 51,792; events = 584) |
| **10** | **Supplemental Figure 7:** Multivariable-adjusted associations of combined sleep, physical activity, and nutrition with MACE risk with excluding poor health individuals (n = 50,787; events = 1,907) |
| **11** | **Supplemental Figure 8:** Multivariable-adjusted associations of combined sleep, physical activity, and nutrition with MACE risk with excluding individuals with an event in the first two years of follow-up (n = 52,946; events = 1,843) |
| **12** | **Supplemental Figure 9:** Multivariable-adjusted associations of combined sleep, physical activity, and nutrition with MACE risk adjusted for BMI (n = 52,744; events = 2,004) |
| **13** | **Supplemental Figure 10:** Multivariable-adjusted associations of combined sleep, physical activity, and nutrition with MACE risk adjusted for sleep characteristics (n = 33,518; events = 1241) |
| **14** | **Supplemental Figure 11:** Multivariable-adjusted associations of combined sleep, physical activity, and nutrition with MACE risk using the proportion of ultra-processed food (n = 39,533; events = 1,462) |
| **15** | **Supplemental Figure 12:** Multivariable-adjusted associations of combined sleep, physical activity, and nutrition with MACE risk adjusted for total energy intake (n = 38,921; events = 1,441) |
| **16** | **Supplementary Methods 1:** Additional study design details |
| **17** | **Supplementary Methods 2**. Wearable behaviour classification methods |
| **20** | **Supplemental Table 1**: Diet quality score index for food-frequency questionnaire dietary data |
| **21** | **Supplemental Table 3**: Covariate definitions |
| **23** | **Supplementary Table 4:** NOVA classification of food groups for 24-hour dietary recall data |
| **24** | **Supplemental Table 6:** STROBE statement |

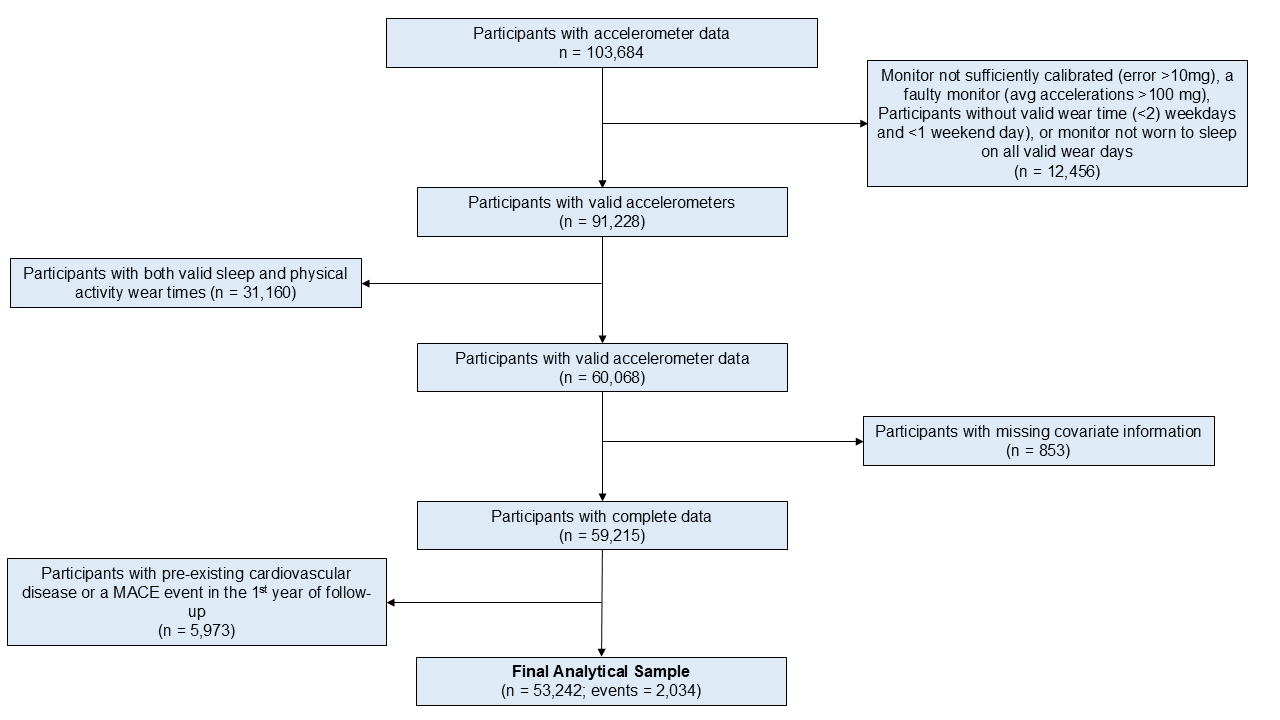

**Supplementary Figure 1.** Participant flow chart

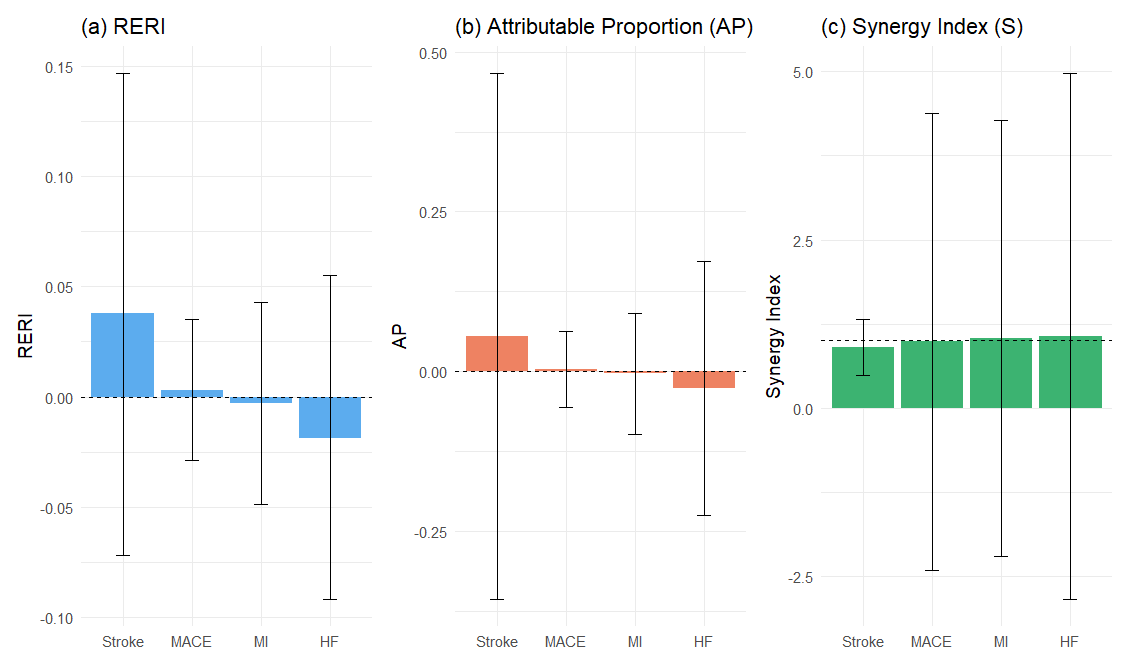

**Supplementary Figure 2:** Synergistic relationship between sleep, physical activity, and nutrition in relation to major adverse cardiovascular events

**Legend**: The figure above shows the individual and interactive model terms of sleep, physical activity, and nutrition for major adverse cardiovascular events (MACE) and its subtypes including stroke, myocardial infarction (MI), and heart failure (HF). To test for interactive and synergistic effects, we calculated the relative excess risk due to interaction (RERI), attributable proportion due to interaction (AP), and the synergistic effects index (S)^1^. These tests provide insight into the contribution of synergistic interactions between exposures, where an RERI or AP of 0 and an S value of 1 denote no interaction effect.

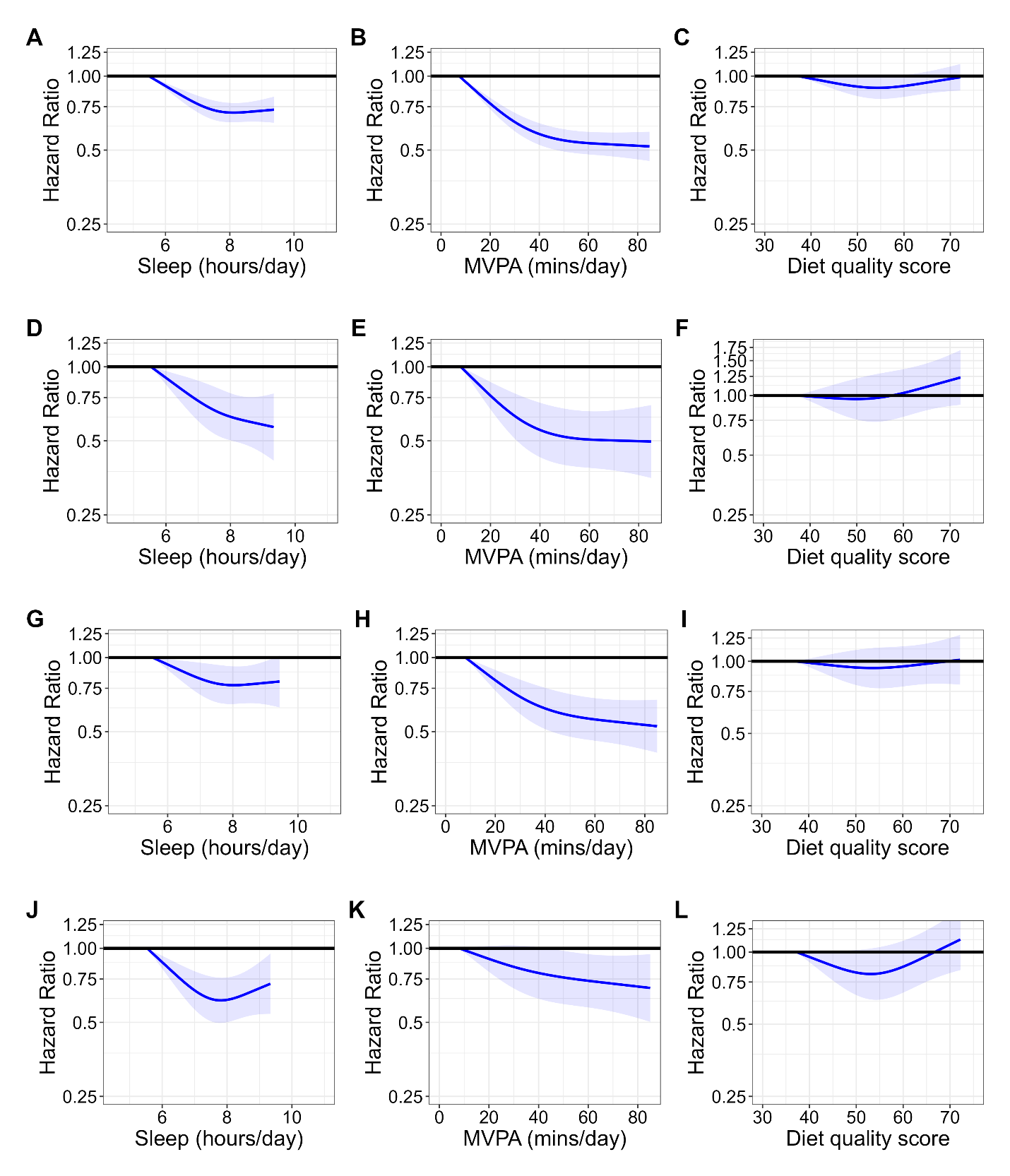

**Supplementary Figure 3:** Dose-response association of sleep, physical activity, and diet with MACE (A-C), heart failure (D-F), myocardial infarction (G-I), and stroke (J-L)

**Legend**: Dose-response plots for sleep, physical activity, and diet from left to right and the rows display with MACE (A-C), heart failure (D-F), myocardial infarction (G-I), and stroke (J-L). Model is adjusted for age, sex, ethnicity, smoking, education, Townsend deprivation index, alcohol, discretionary screen time (time spent watching TV or using the computer outside of work), light intensity physical activity, medication (blood pressure, insulin, and cholesterol), previous diagnosis of cancer, and familial history of CVD and cancer. The reference being the 5th percentile of sleep (5.5 hours/day), physical activity (7.9 minutes/day), and nutrition (37.2 DQS). Participants with a previous diagnosis of major CVD (defined as disease of the circulatory system, arteries, and lymph, excluding hypertension) were excluded from the analysis.

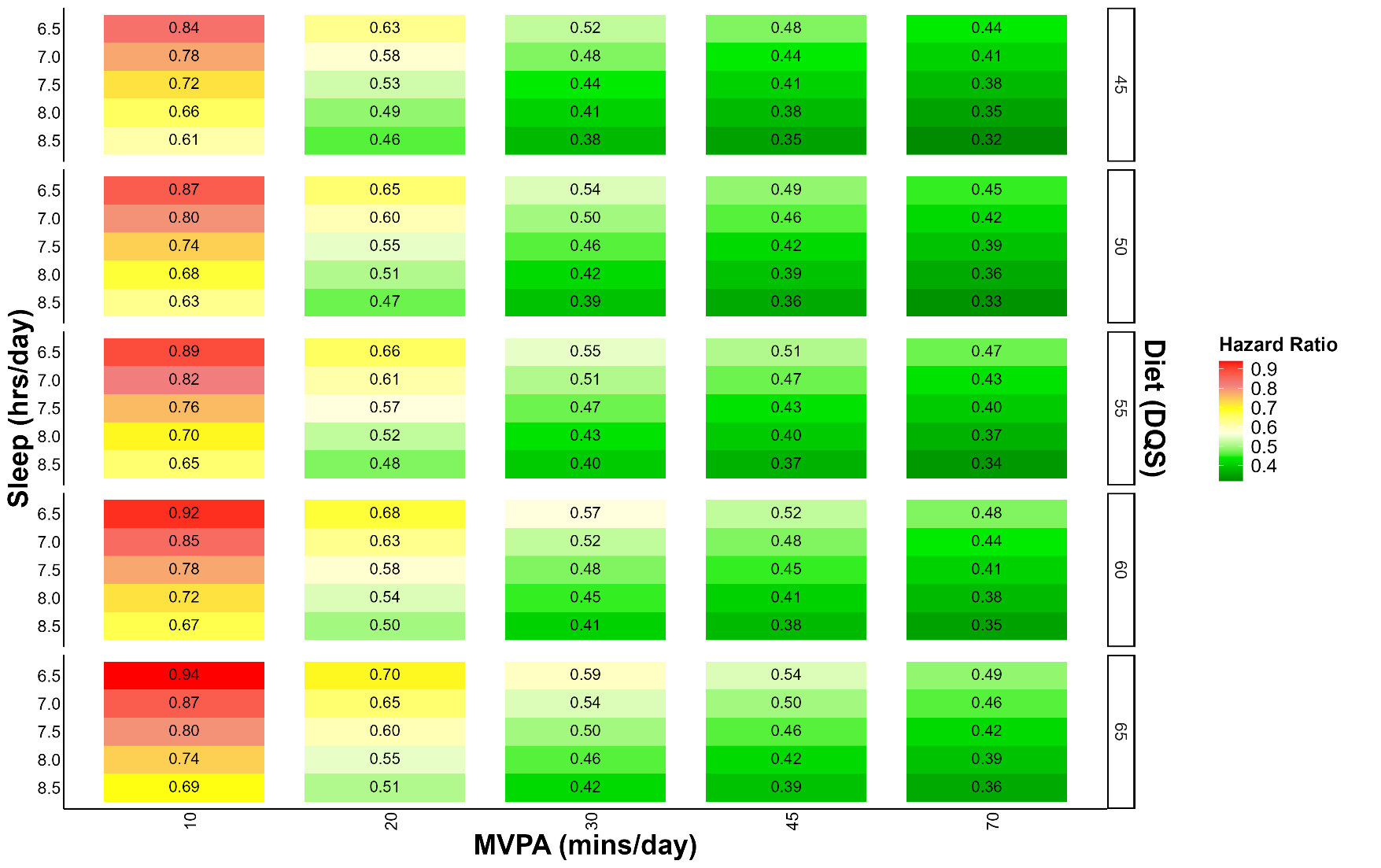

**Supplementary Figure 4**: Multivariable adjusted heart failure risk associated with concurrent variations in sleep, physical activity, and nutrition (n = 51,726; events = 518)

**Legend:** The correlogram displays changes in sleep (hours/day), physical activity (moderate to vigorous intensity (MVPA) minutes/day), and nutrition (Dietary Quality Score (DQS)) and corresponding heart failure risk with the reference being the 5^th^ percentile of sleep (5.5 hours/day), physical activity (7.9 minutes/day), and nutrition (37.2 DQS). Sleep, physical activity, and nutrition are included as independent terms in the model to allow for more granular predictions. Each square on the grid represents the hazard ratio for heart failure associated with a combination of behaviours, as defined by the x-axis (physical activity), y-axis (sleep), and z-axis (nutrition). The colour corresponds to the hazard ratio where red indicates a higher risk of heart failure and green indicates a lower risk of heart failure. Model is adjusted for age, sex, ethnicity, smoking, education, Townsend deprivation index, alcohol, discretionary screen time (time spent watching TV or using the computer outside of work), light intensity physical activity, medication (blood pressure, insulin, and cholesterol), previous diagnosis of cancer, and familial history of CVD and cancer. Participants with a previous diagnosis of major CVD (defined as disease of the circulatory system, arteries, and lymph, excluding hypertension) were excluded from the analysis.

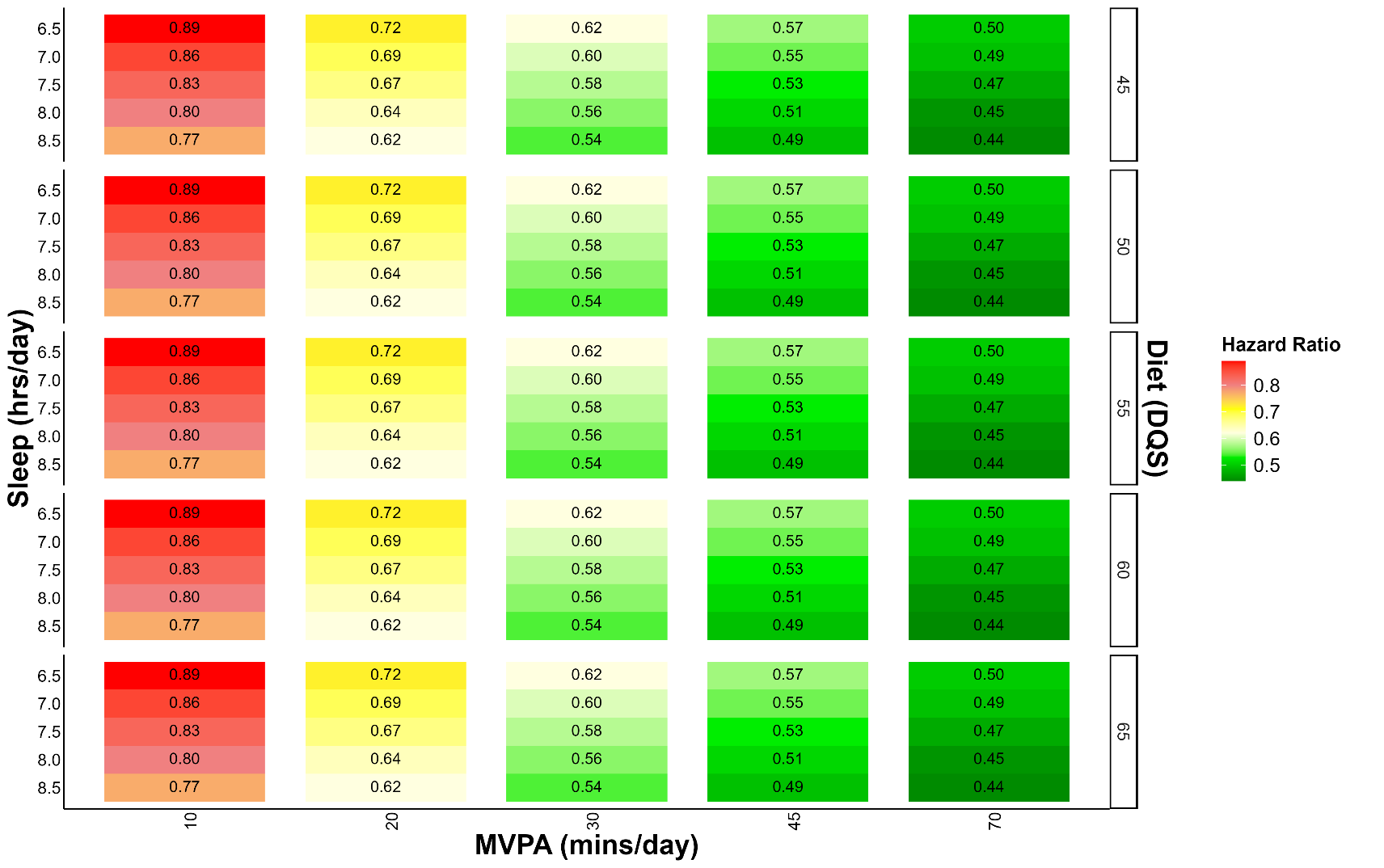

**Supplementary Figure 5**: Multivariable adjusted myocardial infarction risk associated with concurrent variations in sleep, physical activity, and nutrition (n = 52,140; events = 932)

**Legend:** The correlogram displays changes in sleep (hours/day), physical activity (moderate to vigorous intensity (MVPA) minutes/day), and nutrition (Dietary Quality Score (DQS)) and corresponding myocardial infarction risk with the reference being the 5^th^ percentile of sleep (5.5 hours/day), physical activity (7.9 minutes/day), and nutrition (37.2 DQS). Sleep, physical activity, and nutrition are included as independent terms in the model to allow for more granular predictions. Each square on the grid represents the hazard ratio for myocardial infarction associated with a combination of behaviours, as defined by the x-axis (physical activity), y-axis (sleep), and z-axis (nutrition). The colour corresponds to the hazard ratio where red indicates a higher risk of myocardial infarction and green indicates a lower risk of myocardial infarction. Model is adjusted for age, sex, ethnicity, smoking, education, Townsend deprivation index, alcohol, discretionary screen time (time spent watching TV or using the computer outside of work), light intensity physical activity, medication (blood pressure, insulin, and cholesterol), previous diagnosis of cancer, and familial history of CVD and cancer. Participants with a previous diagnosis of major CVD (defined as disease of the circulatory system, arteries, and lymph, excluding hypertension) were excluded from the analysis.

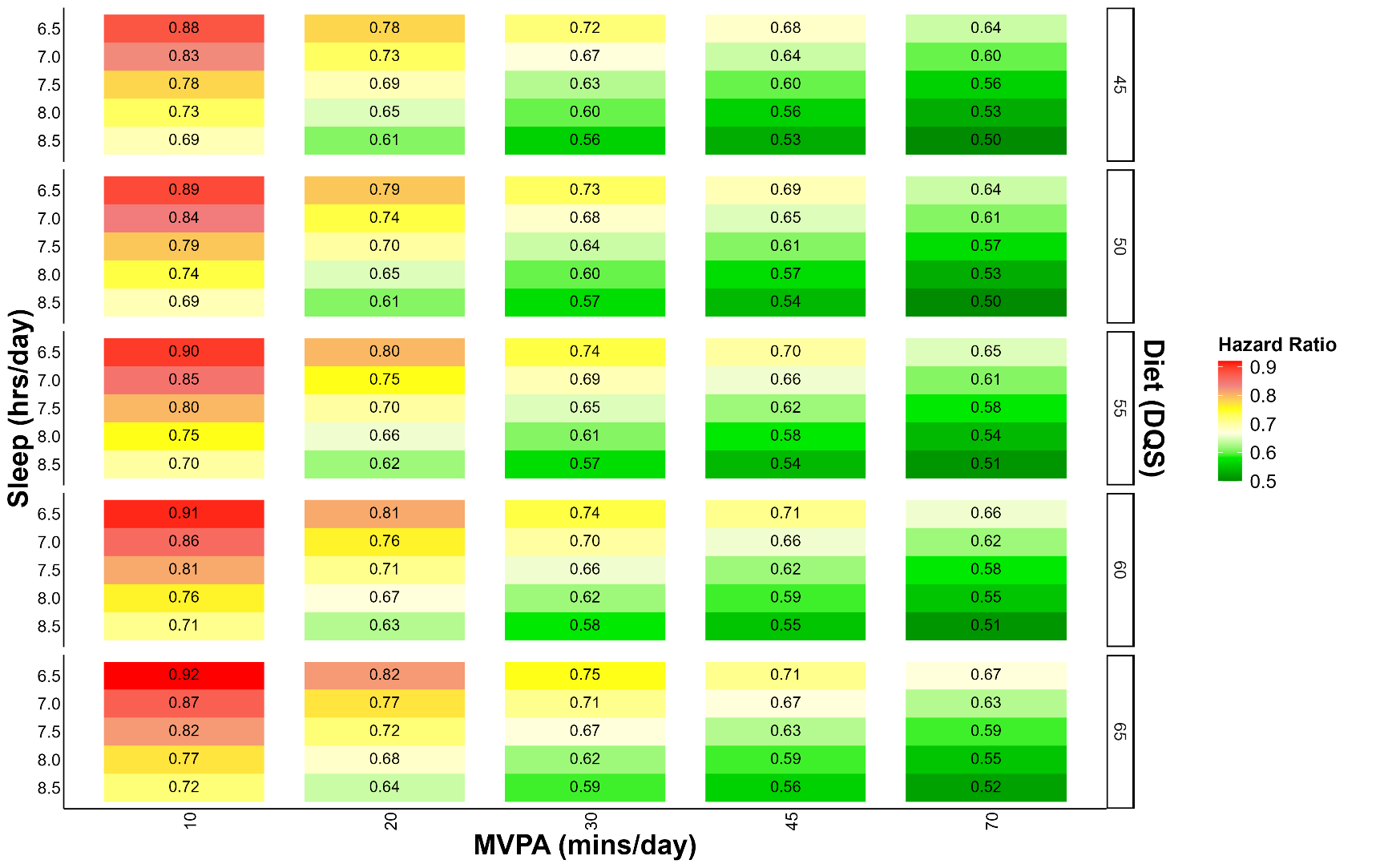

**Supplementary Figure 6**: Multivariable adjusted stroke risk associated with concurrent variations in sleep, physical activity, and nutrition (n = 51,792; events = 584)

**Legend:** The correlogram displays changes in sleep (hours/day), physical activity (moderate to vigorous intensity (MVPA) minutes/day), and nutrition (Dietary Quality Score (DQS)) and corresponding stroke risk with the reference being the 5^th^ percentile of sleep (5.5 hours/day), physical activity (7.9 minutes/day), and nutrition (37.2 DQS). Sleep, physical activity, and nutrition are included as independent terms in the model to allow for more granular predictions. Each square on the grid represents the hazard ratio for stroke associated with a combination of behaviours, as defined by the x-axis (physical activity), y-axis (sleep), and z-axis (nutrition). The colour corresponds to the hazard ratio where red indicates a higher risk of stroke and green indicates a lower risk of stroke. Model is adjusted for age, sex, ethnicity, smoking, education, Townsend deprivation index, alcohol, discretionary screen time (time spent watching TV or using the computer outside of work), light intensity physical activity, medication (blood pressure, insulin, and cholesterol), previous diagnosis of cancer, and familial history of CVD and cancer. Participants with a previous diagnosis of major CVD (defined as disease of the circulatory system, arteries, and lymph, excluding hypertension) were excluded from the analysis.

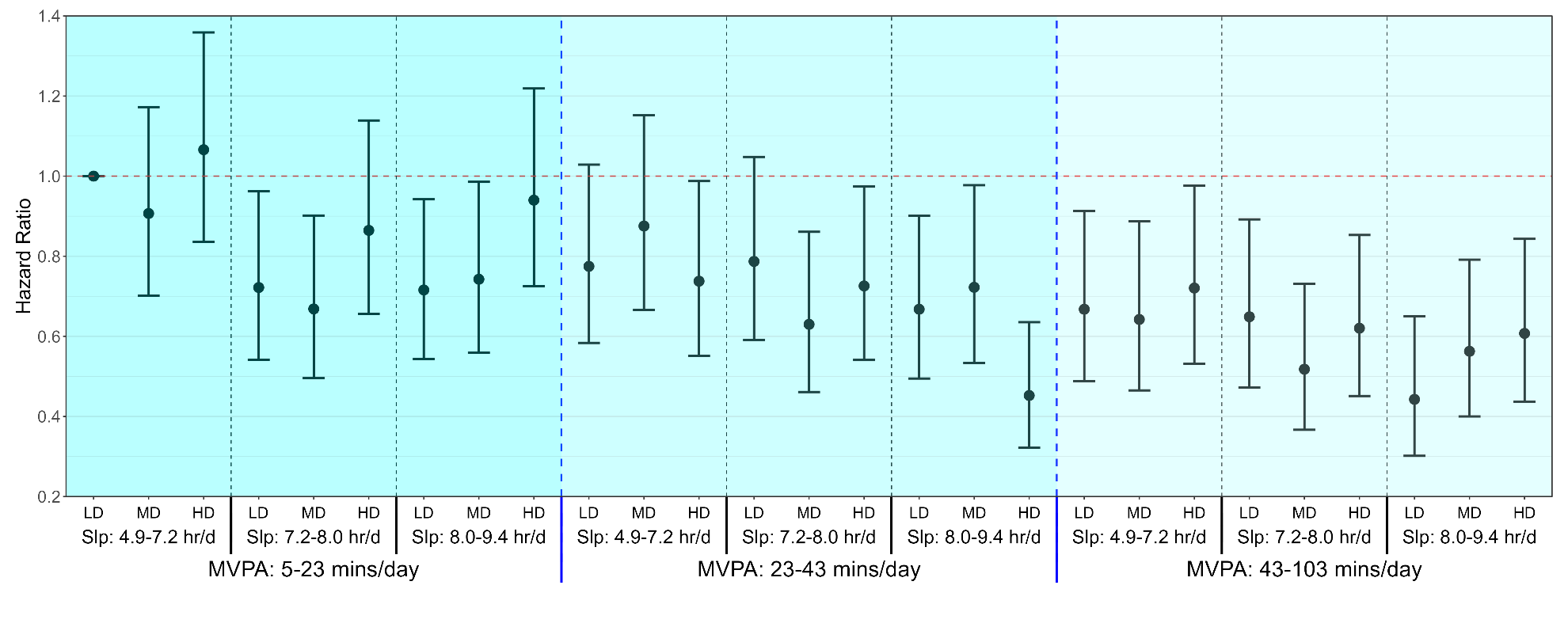

**Supplementary Figure 7.** Multivariable-adjusted associations of combined sleep, physical activity, and nutrition with MACE excluding poor health individuals (n = 50,787; events = 1,907)

**Legend:** Forest plot shows the SPAN associations with MACE after removing those with poor health status including, low BMI (<18.5), current smokers, self-reported poor health, and those with a frailty index score of >3. Model is adjusted for age, sex, ethnicity, smoking, education, Townsend deprivation index, alcohol, discretionary screen time (time spent watching TV or using the computer outside of work), light intensity physical activity, medication (blood pressure, insulin, and cholesterol), previous diagnosis of cancer, and familial history of CVD and cancer. Sleep (hours/day), physical activity (moderate to vigorous intensity (MVPA) minutes/day), and nutrition (Dietary Quality Score (DQS)) were included in the model as a joint term. Participants with a previous diagnosis of major CVD (defined as disease of the circulatory system, arteries, and lymph, excluding hypertension) were excluded from the analysis. The specific ranges for each exposure included sleep duration as 4.8-7.2 hours/day (low), 7·2-8.0 hours/day (medium), and 8.0-9.4 hours/day (high); MVPA measurements as 5-23 minutes/day (low), 23-42 minutes/day (medium), and 42-103 minutes/day (high); and diet quality using the DQS as 32.5-50.0 (low), 50.0-57.5 (medium), and 57.5-72.5 (high). The lowest tertiles for all three exposures (sleep, MVPA and DQS) was considered the reference group. Dashed blue lines separate tertiles MVPA and dashed black lines separate tertiles of sleep. Sleep (Slp); Low Diet Quality (LD); Medium Diet Quality (MD); High Diet Quality (HD).

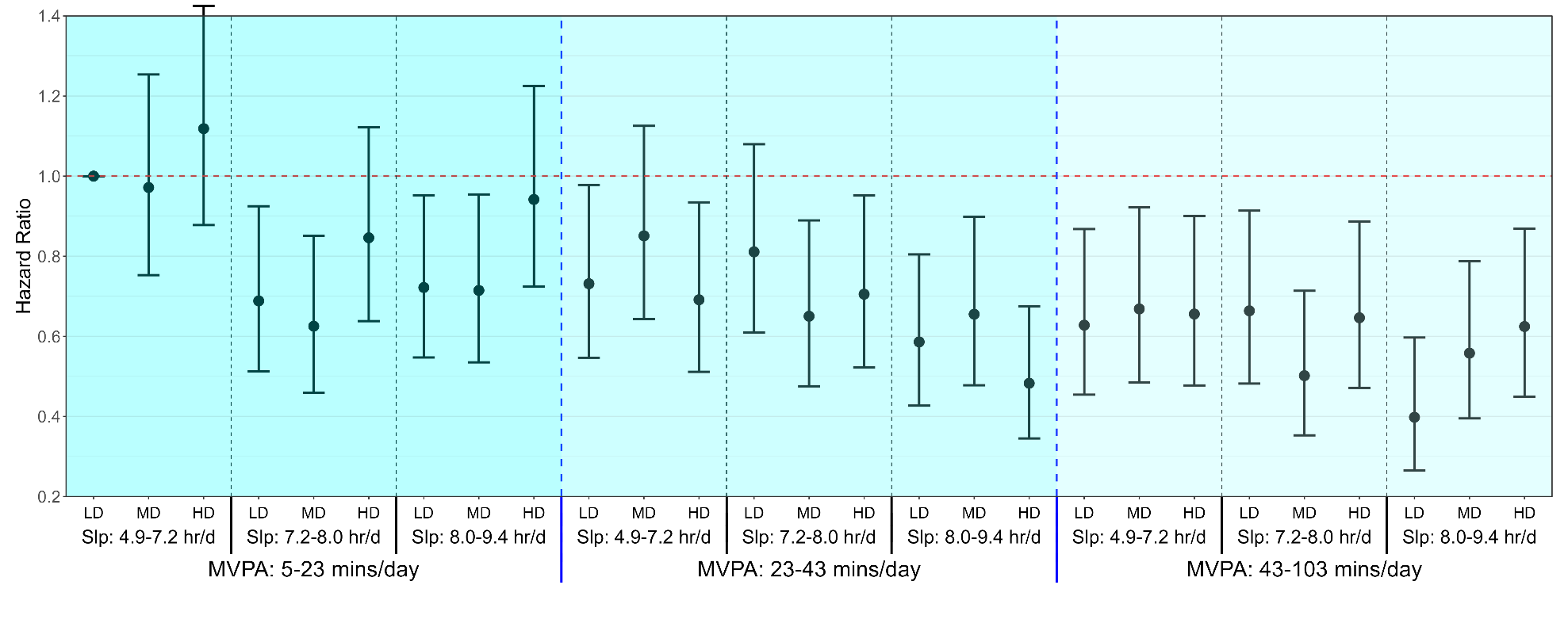

**Supplemental Figure 8**: Multivariable-adjusted associations of combined sleep, physical activity, and nutrition with MACE risk with excluding individuals with an event in the first two years of follow-up (n = 52,946; events = 1,843)

**Legend:** Forest plot shows the SPAN associations with MACE after removing those with an event in the first two years of follow-up. Model is adjusted for age, sex, ethnicity, smoking, education, Townsend deprivation index, alcohol, discretionary screen time (time spent watching TV or using the computer outside of work), light intensity physical activity, medication (blood pressure, insulin, and cholesterol), previous diagnosis of cancer, and familial history of CVD and cancer. Sleep (hours/day), physical activity (moderate to vigorous intensity (MVPA) minutes/day), and nutrition (Dietary Quality Score (DQS)) were included in the model as a joint term. Participants with a previous diagnosis of major CVD (defined as disease of the circulatory system, arteries, and lymph, excluding hypertension) were excluded from the analysis. The specific ranges for each exposure included sleep duration as 4.8-7.2 hours/day (low), 7·2-8.0 hours/day (medium), and 8.0-9.4 hours/day (high); MVPA measurements as 5-23 minutes/day (low), 23-42 minutes/day (medium), and 42-103 minutes/day (high); and diet quality using the DQS as 32.5-50.0 (low), 50.0-57.5 (medium), and 57.5-72.5 (high). The lowest tertiles for all three exposures (sleep, MVPA and DQS) was considered the reference group. Dashed blue lines separate tertiles MVPA and dashed black lines separate tertiles of sleep. Sleep (Slp); Low Diet Quality (LD); Medium Diet Quality (MD); High Diet Quality (HD).

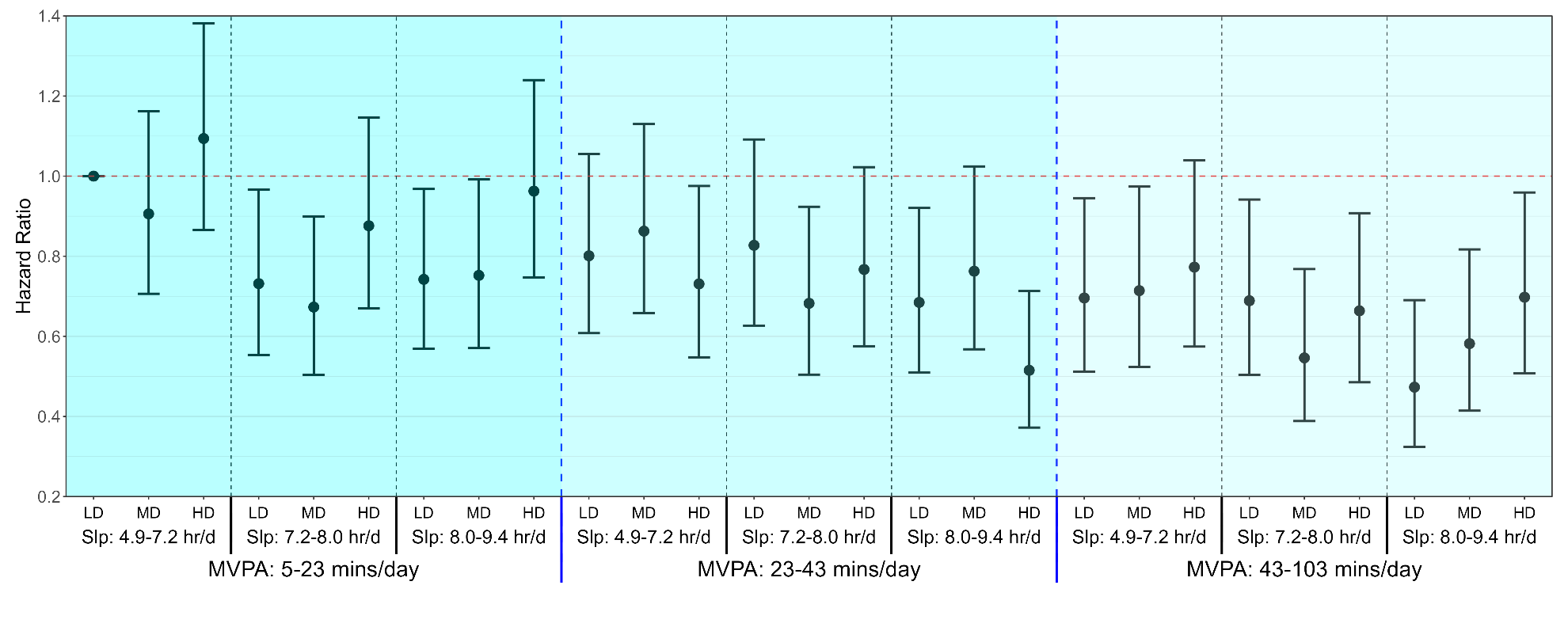

**Supplemental Figure 9:** Multivariable-adjusted associations of combined sleep, physical activity, and nutrition with MACE risk adjusted for BMI (n = 52,744; events = 2,004)

**Legend**: Model is adjusted for age, sex, ethnicity, smoking, education, Townsend deprivation index, alcohol, discretionary screen time (time spent watching TV or using the computer outside of work), light intensity physical activity, medication (blood pressure, insulin, and cholesterol), previous diagnosis of cancer, and familial history of CVD or cancer, and BMI. Sleep (hours/day), physical activity (moderate to vigorous intensity (MVPA) minutes/day), and nutrition (Dietary Quality Score (DQS)) were included in the model as a joint term. Participants with a previous diagnosis of major CVD (defined as disease of the circulatory system, arteries, and lymph, excluding hypertension) were excluded from the analysis. The specific ranges for each exposure included sleep duration as 4.8-7.2 hours/day (low), 7·2-8.0 hours/day (medium), and 8.0-9.4 hours/day (high); MVPA measurements as 5-23 minutes/day (low), 23-42 minutes/day (medium), and 42-103 minutes/day (high); and diet quality using the DQS as 32.5-50.0 (low), 50.0-57.5 (medium), and 57.5-72.5 (high). The lowest tertiles for all three exposures (sleep, MVPA and DQS) was considered the reference group. Dashed blue lines separate tertiles MVPA and dashed black lines separate tertiles of sleep. Sleep (Slp); Low Diet Quality (LD); Medium Diet Quality (MD); High Diet Quality (HD).

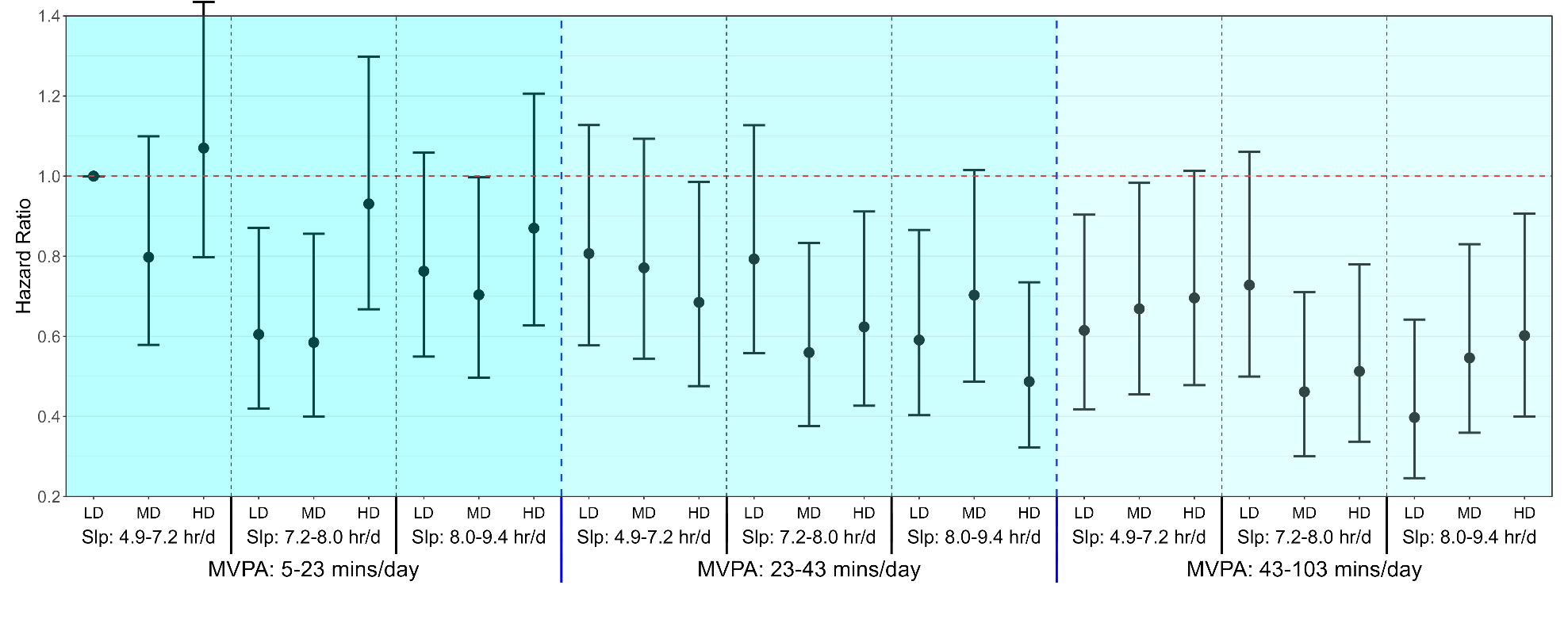

**Supplementary Figure 10:** Multivariable-adjusted associations of combined sleep, physical activity, and nutrition with MACE risk adjusted for sleep characteristics (n = 33,518; events = 1,241)

**Legend**: Model is adjusted for age, sex, ethnicity, smoking, education, Townsend deprivation index, alcohol, discretionary screen time (time spent watching TV or using the computer outside of work), light intensity physical activity, medication (blood pressure, insulin, and cholesterol), previous diagnosis of cancer, and familial history of CVD or cancer, insomnia, snoring, chronotype (morning/evening person), and daytime sleepiness. Sleep (hours/day), physical activity (moderate to vigorous intensity (MVPA) minutes/day), and nutrition (Dietary Quality Score (DQS)) were included in the model as a joint term. Participants with a previous diagnosis of major CVD (defined as disease of the circulatory system, arteries, and lymph, excluding hypertension) were excluded from the analysis. The specific ranges for each exposure included sleep duration as 4.8-7.2 hours/day (low), 7·2-8.0 hours/day (medium), and 8.0-9.4 hours/day (high); MVPA measurements as 5-23 minutes/day (low), 23-42 minutes/day (medium), and 42-103 minutes/day (high); and diet quality using the DQS as 32.5-50.0 (low), 50.0-57.5 (medium), and 57.5-72.5 (high). The lowest tertiles for all three exposures (sleep, MVPA and DQS) was considered the reference group. Dashed blue lines separate tertiles MVPA and dashed black lines separate tertiles of sleep. Sleep (Slp); Low Diet Quality (LD); Medium Diet Quality (MD); High Diet Quality (HD).

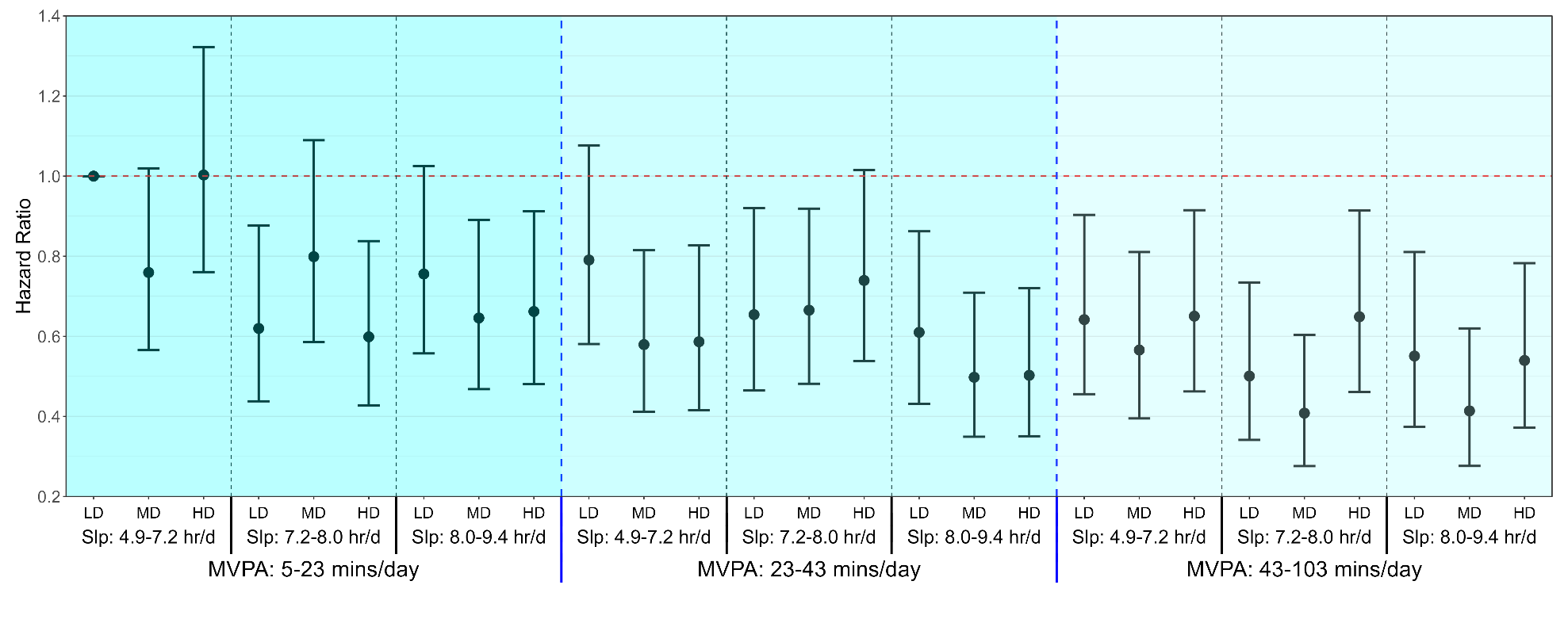

**Supplementary Figure 11:** Multivariable-adjusted associations of combined sleep, physical activity, and nutrition with MACE risk using the proportion of ultra-processed food (n = 39,533; events = 1,462)

**Legend**: Model is adjusted for age, sex, ethnicity, smoking, education, Townsend deprivation index, alcohol, discretionary screen time (time spent watching TV or using the computer outside of work), light intensity physical activity, medication (blood pressure, insulin, and cholesterol), and familial history of CVD and cancer. Sleep (hours/day), physical activity (moderate to vigorous intensity (MVPA) minutes/day), and nutrition (Dietary Quality (UPF%)) were included in the model as a joint term. Diet quality was defined as the percentage of dietary ultra-processed food, where higher diet quality had a lower proportion of ultra-processed food in the diet. The specific ranges for each exposure included sleep duration as 4.8-7.2 hours/day (low), 7.2-8.0 hours/day (medium), and 8.0-9.4 hours/day (high); MVPA measurements as 5-23 minutes/day (low), 23-42 minutes/day (medium), and 42-103 minutes/day (high); and diet quality using the proportion of ultra-processed food as 21.5-100.0 (low), 13.2-21.5 (medium), and 0.0-13.2% (high). Lowest tertiles for all three exposures (sleep, MVPA and diet quality) were considered the reference group. Dashed blue lines separate tertiles MVPA and dashed black lines separate tertiles of sleep. Sleep (Slp); Low Diet Quality (LD); Medium Diet Quality (MD); High Diet Quality (HD).

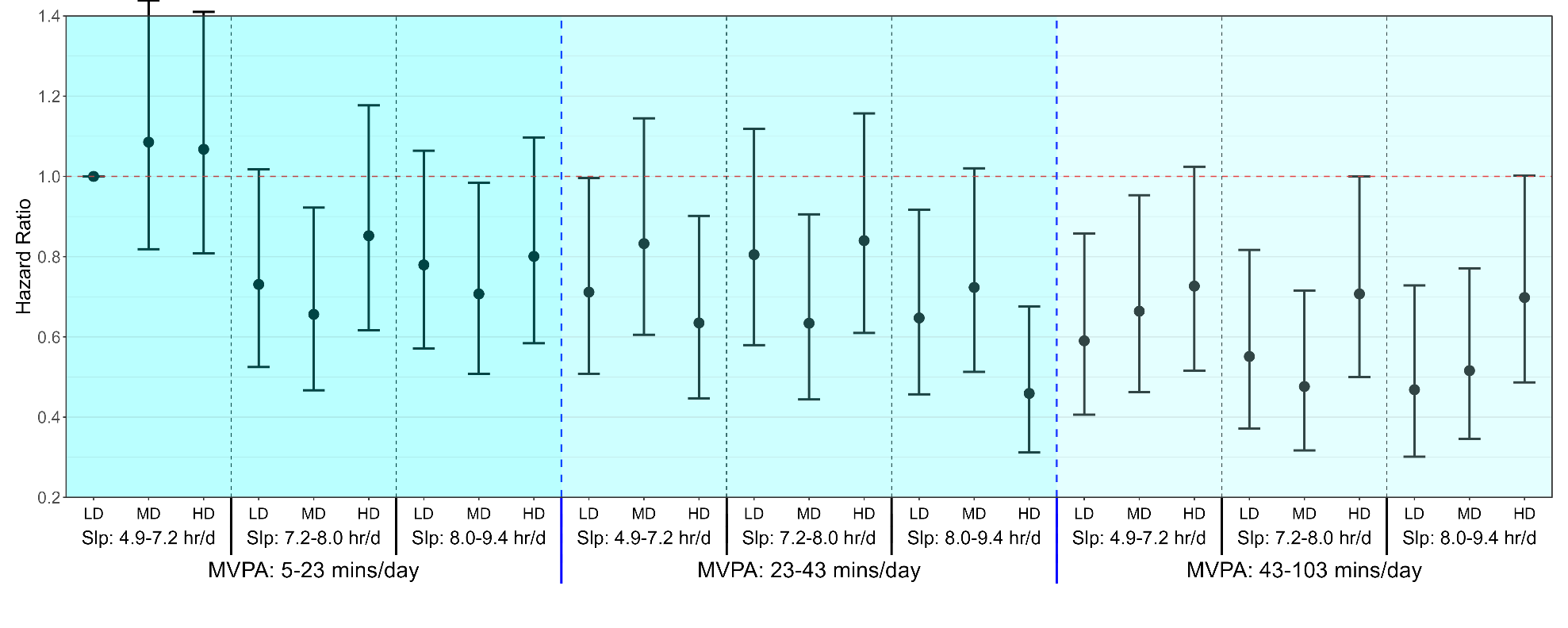

**Supplemental Figure 12**: Multivariable-adjusted associations of combined sleep, physical activity, and nutrition with MACE risk adjusted for total energy intake (n = 38,921; events = 1,441)

**Legend**: Model is adjusted for age, sex, ethnicity, smoking, education, Townsend deprivation index, alcohol, discretionary screen time (time spent watching TV or using the computer outside of work), light intensity physical activity, medication (blood pressure, insulin, and cholesterol), previous diagnosis of cancer, and familial history of CVD or cancer, total energy intake. Sleep (hours/day), physical activity (moderate to vigorous intensity (MVPA) minutes/day), and nutrition (Dietary Quality Score (DQS)) were included in the model as a joint term. Participants with a previous diagnosis of major CVD (defined as disease of the circulatory system, arteries, and lymph, excluding hypertension) were excluded from the analysis. The specific ranges for each exposure included sleep duration as 4.8-7.2 hours/day (low), 7·2-8.0 hours/day (medium), and 8.0-9.4 hours/day (high); MVPA measurements as 5-23 minutes/day (low), 23-42 minutes/day (medium), and 42-103 minutes/day (high); and diet quality using the DQS as 32.5-50.0 (low), 50.0-57.5 (medium), and 57.5-72.5 (high). The lowest tertiles for all three exposures (sleep, MVPA and DQS) was considered the reference group. Dashed blue lines separate tertiles MVPA and dashed black lines separate tertiles of sleep. Sleep (Slp); Low Diet Quality (LD); Medium Diet Quality (MD); High Diet Quality (HD).

**Supplementary** **Methods 1:** Additional study design details

**Study Sample and Design**

This study analyzed the data from the UK Biobank which is an ongoing prospective cohort study comprised of adults aged 40-69 at baseline (2006-2010)^2^. Participants provided informed consent and ethical approval was provided by the UK’s National Health Service, National Research Ethics Service (Ref80 11/NW/0382). Deaths were ascertained through linkage with the National Health Service Digital of England and Wales or the National Health Service Central Register and National Records of Scotland. Censoring for England, Wales, and Scotland was up to November 30th, 2022.

Hospital inpatient data was ascertained through linkage with the National Health Service Digital for England, the Information and Statistics Division for Scotland, and Secure Anonymized Information Linkage for Wales. Censoring for England and Scotland was up to October 31st, 2022 and August 31st, 2022, respectively. Censoring for Wales was up to May 31st, 2022. Cancer data linkage was obtained through national cancer registries. For England and Wales, cancer diagnosis data were followed up through 31 December 2020 and 31 December 2016 respectively, and were provided by NHS England^3^. For Scotland, cancer diagnosis data were followed up through 30 November 2021 and provided by the National Records of Scotland^3^.

Between 2013 and 2015 (median 5.5 years after the baseline measurements), 103,684 UK Biobank participants wore a wrist-worn accelerometer for 7 days^4^. The accelerometers were calibrated before being mailed to the individuals. During the initial processing stage, we excluded participants who if no sleep data was recorded, the accelerometer was poorly calibrated (>10 milli-gravitational units (mg)), or a faulty accelerometer was distributed (>100mg)^5-8^. We excluded participants with missing covariates and insufficient valid wear days. Monitoring days were considered valid if wear time was greater than 16 hours. To be included in the analysis, participants were required to have at least three valid monitoring days, with at least one of those days being a weekend day. We excluded participants who reported that they could not walk^5-8^.

**Supplementary Methods 2**. Wearable behaviour classification methods

**Sleep and non-wear time**

The non-wear time was determined using a previously validated algorithm that uses wrist tilt angle to determine non-wear with 86-95% accuracy^9^. No values were imputed for non-wear time. Sleep was defined as the average daily duration of sleep (hours/day) as calculated using a validated algorithm based on relative changes in wrist tilt angle between successive 5-second windows^10^. For each interval of 5 seconds, the average of the estimated wrist tilt angle was calculated and a rolling 5-minute median served as an input for the algorithm to identify sleep onset and sleep offset, and then time spent asleep within this timeframe^9,10^.

**Two-stage random forest physical activity intensity and posture classification**

Incidental physical activity was classified using a validated two-stage random forest activity classifier that first classifies each 10 second window (epoch) as sedentary (lying or sitting still), stationary plus (active sitting, standing still, active standing), walking, or running (**Diagram A**)^9-11^. These activities were then classified into one of four activities including: sedentary, light, moderate, and vigorous. Walking activities (gardening, active commuting, etc) were classified by normalized gravitational units (g) where <100 milli g were classified as light intensity (<3 METs), ≥100 milli g and <400 milli g were considered moderate intensity physical activity (≥3 to <6 METs), and ≥400 milli g were considered vigorous-intensity PA (≥6 METs)^11^. All windows classified as running/high energetic activity were classified as vigorous-intensity physical activity (≥ 6 METs)^5,7,11^. A major advantage of this classification approach is the lower risk of possible misclassification of sporadic high-accelerations that may occur during certain stationary light activities (e.g., dishwashing)^12-14^.

**Physical Activity Classification Scheme (Diagram A)**

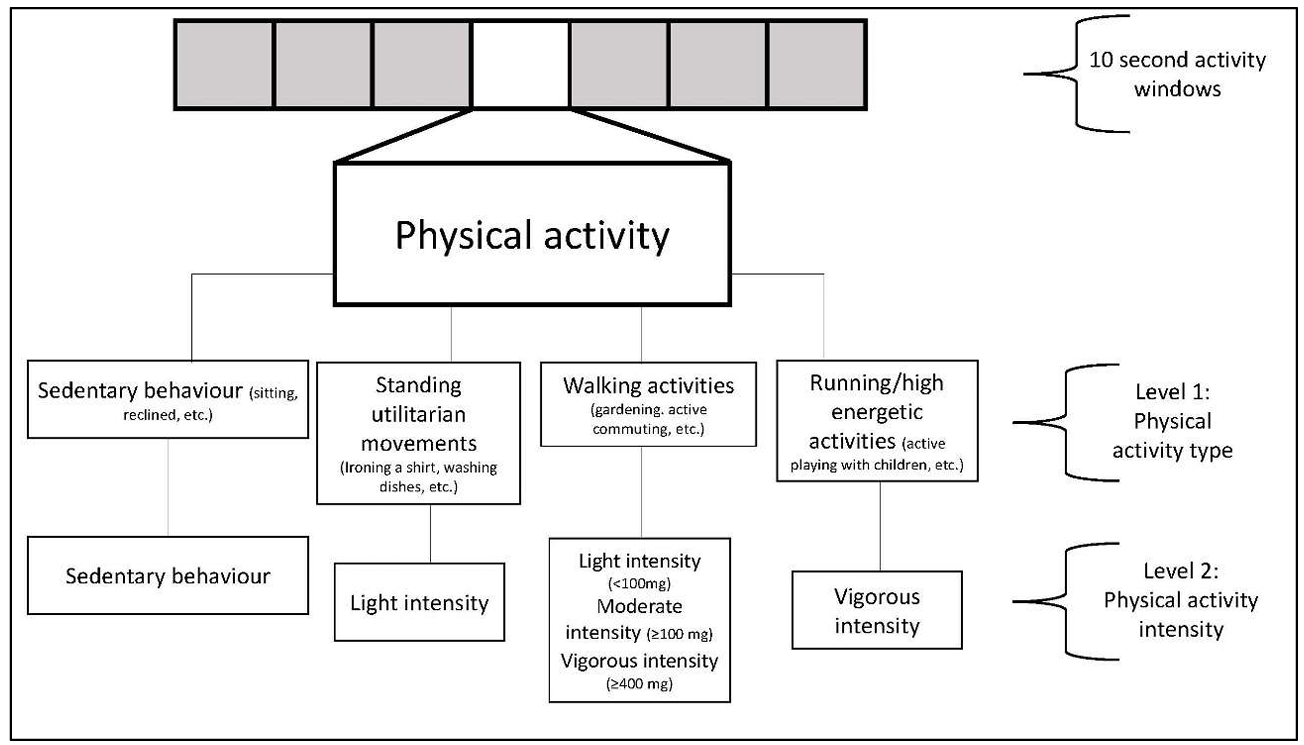

**Physical Activity Classification Performance**

The performance of this physical activity classification scheme was tested in an independent sample of 102 adults from the US^15^ and Australia^16^. This data includes direct observation measurement of 105,767 activity samples from structured and free-living activities (17,627 minutes) which were used to test the robustness and generalisability of the two-stage activity and intensity classifier. The data was collected from participant-worn or researcher-held Go-Pro video recordings. All data was imported into Noldus Observer XT software for continuous video coding. The direct observation coding generated continuous physical activity codes corresponding to the start and finish of each movement. These coded movements were then compared against the accelerometer data using the available time-stamp information. The below table includes the performance metrics across activities. Interobserver reliability was assessed by dual coding. The intraclass correlation coefficient for coding activities was 0.912 (0.866-0.942). The performance in metrics and confusion matrix for activity classification is shown below.

**Classifier Performance Metrics for Intensity in US and Australian Adults**

|  | Sensitivity | Specificity | Precision | F-score | Overall Accuracy | Weighted Kappa | Overall F-score |
| --- | --- | --- | --- | --- | --- | --- | --- |
| Sedentary | 86.5 | 93.7 | 90.5 | 88.5 |  |  |  |
| Light | 71.2 | 89.4 | 55.8 | 62.6 |  |  |  |
| Moderate | 85.4 | 96.6 | 92.7 | 88.9 |  |  |  |
| Vigorous | 95.4 | 99.4 | 94.6 | 95.0 |  |  |  |
|  |  |  |  |  | **84.6** | 0.78 | 83.8 |

Rows= ground truth; columns=predictions; bold=correct classification; all activities were free-living or simulated free-living activities.

**Confusion Matrix for Activity Classification in US and Australian Adults**

|  | Sedentary | Light | Moderate | Vigorous |
| --- | --- | --- | --- | --- |
| Sedentary | **36,904** | 5,232 | 508 | 2 |
| Light | 3,120 | **11,712** | 1,612 | 17 |
| Moderate | 502 | 4,016 | **29,528** | 526 |
| Vigorous | 226 | 17 | 214 | **9,470** |

Rows= ground truth; columns=predictions; bold=correct classification; all activities were free-living or simulated free-living activities.

**Supplementary Table 1.** Diet quality score index for food-frequency questionnaire dietary data

| **Food components** | **UK Biobank field ID** | **Amount per serving** | **Criteria for maximum score (10)** | **Criteria for minimum score (0)** |
| --- | --- | --- | --- | --- |
| Fruit | 1309 (pieces fresh fruit/day) 1319 (pieces dried fruit/day) | 1309 – 1 piece  1319 – 5 pieces | ≥3 servings/day | 0 servings/day |
| Vegetable | 1289 (tablespoons cooked vegetables/day)  1299 (salad/raw vegetables/day) | 3 heaped tablespoons | ≥3 servings/day | 0 servings/day |
| Whole grains | 1438, 1448 (wholemeal/wholegrain bread slices/week)  1458, 1468 (bran/oat/muesli cereal) | 1438/1448 – 1 slice/day 1458/1468 – 1 bowl/day | ≥3 servings/day | 0 servings/day |
| Fish | 1329 (oily fish/week)  1339 (non-oily fish/week) | Once/week | ≥2 servings/week | 0 servings/week |
| Dairy | 1408 (cheese/week)  1418 (milk type) | 1408 – 1 piece/day  1418 – 1 glass/day if consumption of any type of milk | ≥2 servings/day | 0 servings/day |
| Vegetable oils | 1428 (Flora Pro-Active/Benecol spread)  2654 (Flora Pro-Active/Benecol, soft margarine -, olive oil based -, polyunsaturated/sunflower oil based -, other low/reduced fat spread)  1438 (bread slices/week) | 1 serving/day if in combination with eating at least 2 slices of bread (ID 1438) | ≥2 servings/day | 0 servings/day |
| Refined grains | 1438, 1448 (white, brown, other bread slices/week) 1458, 1468 (biscuit, other cereals/week) | 1438/1448 – 1 slice/day 1458/1468 – 1 bowl/day | 0 servings/day | >2 servings/day |
| Processed meats | 1349 (processed meat/week or daily)  3680 (age when last ate meat) | 1349 – 1 piece/day  3680 – 0 pieces/day if indicated having never eaten meat | 0 serving/week | >1 serving/week |
| Unprocessed red meats | 1369 (beef/week or day)  1379 (lamb or mutton/week or day)  1389 (pork/week or day)  3680 (age when last ate meat) | 1359-1389 – once/week 3680 – 0 pieces/day if indicated having never eaten meat | 0 serving/week | >2 serving/week |
| Sugar-sweetened beverages | 6144 (never consumes drinks containing sugar) | 0 servings | Don’t drink | Drink |

Diet quality score information is adapted from previously established work by Zhuang *et al*. Diabetes Care^17^. Intermediate intake for each dietary component were scored relative to minimum to maximum intake of each food component. The formula for intermediate intakes of adequacy components is described as: component score = (maximum score / (Amax - Amin))*(X - Amin) and for moderate components (refined grains, processed meat, and unprocessed red meat) component score = (maximum score - maximum score / (Amax - Amin))*(X - Amin). The food frequency questionnaire demonstrated moderate reproducibility for food groups (Intraclass Correlation Coefficient (ICC): 0.48-0.66) and modest agreement with alternative dietary intake measures from the 24-hour recall (ICC: 0.38-0.63). This level of agreement and reproducibility is comparable to previous prospective observational studies^18-20^. The food frequency questionnaire has also been validated against the 24-hour dietary recall using objective biomarkers as the standard^21^.

**Supplementary Table 2.** Covariate Definitions.

| **Variable** | **Definition** | **UK Biobank field ID (if applicable)** |
| --- | --- | --- |
| Age | Categorical (4) | 34, 52, accelerometer date-timestamp |
| Sex | Female/Male | 31 |
| Ethnicity | White/Others | 21000 |
| Education | College/University; A/AS level; O levels; CSE; NVQ/HND/HNC; other | 6138 |
| Smoking status | Never, past, current | 20116 |
| Alcohol consumption | Units/day | 20403 |
| Light intensity physical activity | Standing utilitarian movements, slow walking (<3 METs) | Derived from accelerometer data |
| Discretionary screen-time | Time spent/day watching TV and using a computer outside of work | 1070, 1080 |
| Townsend deprivation | Categorical (5) | 22189 |
| Use of cholesterol medication | Yes/No | 6177, 6153 |
| Use of blood pressure medication | Yes/No | 6177, 6153 |
| Use of diabetes medication | Yes/No | 6177, 6153 |
| Previous CVD | Identified by self-report and cancer registry. Defined as disease of the circulatory system, arteries, and lymph, excluding hypertension | 20002, 41270 |
| Previous cancer | Identified by self-report and hospitalisation | 20001, 100092 |
| Familial history of CVD | Self-reporter mother of father diagnosed with heart disease or stroke | 20107, 20110 |
| Familial history of cancer | Self-reporter mother of father diagnosed with cancer | 20107, 20110 |
| High frailty scale | Categorical (yes/no); high frailty indicates a score of ≥3 on a 0 to 5 | 2306, 120107, 2624, 1011, 3637, 991, 971, 924, 46, 47 |
| Body mass index | Continuous; kilogram/meter^2^ | 23104 |
| Total energy intake | Continuous, kcal/day | 26002 |
| Morning/evening person (chronotype) | Categorical (definitely a ‘morning’ person; more a ‘morning’ person than ‘evening’ person; more an ‘evening’ person than ‘morning’ person; definitely an ‘evening’ person) | 1180 |
| Insomnia | Categorical (never/rarely; sometimes; usually) | 1200 |
| Snoring | Categorical (yes/no) | 1210 |
| Daytime sleepiness | Categorical (never/rarely; sometimes; often) | 1220 |

Additional detail is available online at https://biobank.ndph.ox.ac.uk/showcase/.

**Supplementary Table 3.** NOVA classification of food groups for 24-hour dietary recall data.

| **NOVA classification level** | **UK Biobank field ID (if applicable)** |
| --- | --- |
| Level four | Added sugars and preserves (26064), Animal fat spread lower fat (26062), Animal fat spread normal (26063), Biscuit cereal (26075), Biscuits (26068), Bran cereal (26076), Breaded/battered chicken (26069), Breaded/battered fish (26070), Chocolate confectionery (26080), Cream (26154), Fried/roast potatoes (26119), Low/non sugar sugar-sweetened beverages (26126), Mashed potatoes (26120), Meat substitutes - soy (26137), Meat substitutes - vegetarian (26145), Milk-based and powdered drinks (26087), Milk-dairy desserts (26084), Mixed bread brown and seeded (26071), Muesli (26105), Nut-based spreads (26106), Other cereal (sugar) (26079), Other desserts and cakes and pastries (26085), Other sweets (26140), Pizza (26116), Plant-based spread lower fat (26111), Plant-based spread normal (26112), Processed meat (26122), Samosa, pakora (26128), Sauces and condiments (high fat) (26129), Sauces and condiments (low fat) (26130), Savoury crackers (26083), Savoury snacks (26134), Soy desserts and yogurt (26086), Sugar-sweetened beverages and other sugary drinks (26127), Sushi (26139), Vegetable dips (26144) |
| Level three | High fat cheese (26099), Medium and low fat cheese (26103), White fish and tinned tuna (26149), White bread (26073), Wholemeal bread (26074), Other bread (26072) |
| Level two | Grain dishes - added fat (26097), Olive oil (drizzling/dunking) (26110) |
| Level one | Allium vegetables (26065), Apples and pears (26089), Beef (26066), Berries (26090), Citrus (26091), Coffee, caffeinated (26081), Coffee, decaffeinated (26082), Dried fruit (26092), Egg and egg dishes (26088), Fruit juice (26095), Green leafy/cabbages (26098), Lamb (26100), Legumes and pulses (26101), Oat cereal (non sugar) (26077), Oat cereal (sugar) (26078), Low fat yogurt (26102), Full fat yogurt (26096), Oily fish (26109), Other fruit (26093), Other meat, offal (26104), Other vegetables, including mushrooms, fruiting and mixed vegetables (26146), Peas and sweetcorn (26115), Pork (26117), Potatoes and sweet potatoes (baked/boiled) (26118), Poultry (26121), Raw salad (26123), Root vegetables (26125), Salted nuts and seeds (26108), Semi skimmed milk (26131), Rice/oat milk (26124), Shellfish (26132), Skimmed milk and cholesterol-lowering milk (26133), Soups (26135), Soy milk (26136), Stewed fruit (26094), Tea (26141), Tea, decaffeinated (26142), Tomatoes (26143), Unsalted nuts and seeds (26107), White pasta and rice (26113), Whole milk (26150), Wholemeal pasta, brown rice and other wholegrains (26114) |

From 2009-2012, dietary data was also collected using 1-4 separate 24-hour dietary recalls for a subgroup of participants (n = 211,031)^22^. Additional detail on reproducibility and agreement between FFQ and the 24-hour dietary recall has been published elsewhere^23,24^. Food groups in each NOVA classification level were reported as the average weight (gram/day) from the 24-hour dietary recalls. Ultra-processed food intake was defined as the percentage of level four NOVA food groups relative to the average reported total food weight. All food categories and the definition of ultra-processed food intake were determined using a previously established method^25,26^.

**Supplementary Table 4.** STROBE**.**

|  | | Item No | Recommendation | Page No |
| --- | --- | --- | --- | --- |
| **Title and abstract** | | 1 | (*a*) Indicate the study’s design with a commonly used term in the title or the abstract |  |
|  |  |  | (*b*) Provide in the abstract an informative and balanced summary of what was done and what was found | 1-2 |
| Introduction | | | | |
| Background/rationale | | 2 | Explain the scientific background and rationale for the investigation being reported | 3-4 |
| Objectives | | 3 | State specific objectives, including any prespecified hypotheses | 4 |
| Methods | | | | |
| Study design | | 4 | Present key elements of study design early in the paper | 4 |
| Setting | | 5 | Describe the setting, locations, and relevant dates, including periods of recruitment, exposure, follow-up, and data collection | 4-5 |
| Participants | | 6 | (*a*) Give the eligibility criteria, and the sources and methods of selection of participants. Describe methods of follow-up | 4-5 |
|  |  |  | (*b*) For matched studies, give matching criteria and number of exposed and unexposed | - |
| Variables | | 7 | Clearly define all outcomes, exposures, predictors, potential confounders, and effect modifiers. Give diagnostic criteria, if applicable | 5-6 |
| Data sources/ measurement | | 8* | For each variable of interest, give sources of data and details of methods of assessment (measurement). Describe comparability of assessment methods if there is more than one group | 4-6 |
| Bias | | 9 | Describe any efforts to address potential sources of bias | 7-8 |
| Study size | | 10 | Explain how the study size was arrived at | Supplementary Figure 1 |
| Quantitative variables | | 11 | Explain how quantitative variables were handled in the analyses. If applicable, describe which groupings were chosen and why | 5-7 |
| Statistical methods | | 12 | (*a*) Describe all statistical methods, including those used to control for confounding |  |
|  |  |  | (*b*) Describe any methods used to examine subgroups and interactions |  |
|  |  |  | (*c*) Explain how missing data were addressed | 5-8 |
|  |  |  | (*d*) If applicable, explain how loss to follow-up was addressed |  |
|  |  |  | (*e*) Describe any sensitivity analyses |  |
| Results | | | |  |
| Participants | | 13* | (a) Report numbers of individuals at each stage of study—eg numbers potentially eligible, examined for eligibility, confirmed eligible, included in the study, completing follow-up, and analysed | Supplementary Figure 1 |
|  |  |  | (b) Give reasons for non-participation at each stage | Supplemental Figure 1 |
|  |  |  | (c) Consider use of a flow diagram | Supplemental Figure 1 |
| Descriptive data | | 14* | (a) Give characteristics of study participants (eg demographic, clinical, social) and information on exposures and potential confounders | Table 1 |
|  |  |  | (b) Indicate number of participants with missing data for each variable of interest | Supplemental Figure 1 |
|  |  |  | (c) Summarise follow-up time (eg, average and total amount) | Table 1 |
| Outcome data | | 15* | Report numbers of outcome events or summary measures over time |  |
| Main results | 16 | (*a*) Give unadjusted estimates and, if applicable, confounder-adjusted estimates and their precision (eg, 95% confidence interval). Make clear which confounders were adjusted for and why they were included | | 9-11 |
|  |  | (*b*) Report category boundaries when continuous variables were categorized | | 9-11 |
|  |  | (*c*) If relevant, consider translating estimates of relative risk into absolute risk for a meaningful time period | | - |
| Other analyses | 17 | Report other analyses done—eg analyses of subgroups and interactions, and sensitivity analyses | | 11 |
| Discussion | | | | |
| Key results | 18 | Summarise key results with reference to study objectives | | 12 |
| Limitations | 19 | Discuss limitations of the study, taking into account sources of potential bias or imprecision. Discuss both direction and magnitude of any potential bias | | 15-16 |
| Interpretation | 20 | Give a cautious overall interpretation of results considering objectives, limitations, multiplicity of analyses, results from similar studies, and other relevant evidence | | 16 |
| Generalisability | 21 | Discuss the generalisability (external validity) of the study results | | 16 |
| Other information | | | | |
| Funding | 22 | Give the source of funding and the role of the funders for the present study and, if applicable, for the original study on which the present article is based | | 17 |

*Give information separately for exposed and unexposed groups.

**Note:** An Explanation and Elaboration article discusses each checklist item and gives methodological background and published examples of transparent reporting. The STROBE checklist is best used in conjunction with this article (freely available on the websites of PLoS Medicine at http://www.plosmedicine.org/, Annals of Internal Medicine at http://www.annals.org/, and Epidemiology at http://www.epidem.com/). Information on the STROBE Initiative is available at http://www.strobe-statement.org.
